# The STEMRI trial: magnetic resonance spectroscopy imaging can define tumor areas enriched in glioblastoma-initiating cells

**DOI:** 10.1101/2023.01.18.23284651

**Authors:** Anthony Lemarié, Vincent Lubrano, Caroline Delmas, Amélie Lusque, Juan-Pablo Cerapio, Marion Perrier, Aurore Siegfried, Florent Arnauduc, Yvan Nicaise, Perrine Dahan, Thomas Filleron, Muriel Mounier, Christine Toulas, Elizabeth Cohen-Jonathan Moyal

## Abstract

**BACKGROUND:** Glioblastoma (GB) gold standard treatment combines maximally-safe surgical resection of the contrast-enhanced (CE) central tumor area, as defined by MRI, and chemo-radiotherapy. However, most patients relapse within one year in non-CE peritumoral FLAIR regions. Spectroscopy MRI (MRSI) can discriminate metabolic tumor areas with higher recurrence potential. We showed that regions with Choline/N-acetyl-aspartate index >2 (CNI+) were predictive of relapse sites post-radiotherapy in CE and FLAIR areas. As relapses are mainly imputed to a subpopulation of aggressive and resistant tumor stem-like cells, called GB-initiating cells (GIC), this suggests that CNI+ areas might be enriched in GIC.

**METHODS:** We conducted a prospective trial in 16 eligible GB patients subjected to preoperative MRSI/MRI and subsequent surgery/chemo-radiotherapy to investigate GIC enrichment of CNI+ versus CNI− areas, based on biopsies in CE and FLAIR. We combined in vitro/vivo biological characterization of biopsies and derived GIC lines with biopsy RNAseq analyses.

**RESULTS:** Biopsy characterization by FACS and RNAseq revealed that FLAIR/CNI+ areas showed an enrichment in GIC-population and in stem-related gene signature, but also in pathways related to DNA repair, adhesion/migration and mitochondrial bioenergetics. More, FLAIR/CNI+ samples gave rise to GIC-enriched neurospheres faster than CNI− counterparts. Finally, parameters assessing Biopsy GIC Content and Time to Neurosphere formation in FLAIR/CNI+ areas were associated with worse patient outcome.

**CONCLUSION:** Preoperative MRI/MRSI combination would certainly allow better resection and targeting of CNI+ areas in FLAIR, as their GIC-enrichment can predict worse outcome in GB patients.

**TRIAL REGISTRATION:** ClinicalTrials.gov NCT01872221.

**FUNDING:** RITC (RECF1929), GRICR and Plan Cancer 2016 (HTE).

## INTRODUCTION

Glioblastoma (GB, Grade IV glioma) is the most frequent and aggressive malignant primary adult brain cancer. Surgical resection followed by chemo-radiotherapy (30×2Gy and Temozolomide) is the current standard of care. However, prognosis remains extremely poor, with a median survival of ~15 months (1). Recurrences may be notably explained by the major cellular heterogeneity of these tumors which contain GB-initiating cells (GIC) (2). GIC are characterized by their unlimited self-renew ability, their stem markers overexpression (Sox2, CD133, Nestin, Olig2, A2B5, ITGA6…), their multipotent aptitude to differentiate into neural lineages, their invasion capacity, their localization in perivascular niches, their strong chemo/radioresistance and an increased tumorigenic potential (2, 3). Consequently, GIC targeting is a strong therapeutic challenge in GB management.

The tumor heterogeneity can be visualized and mapped by multimodal imaging including multi-modal Magnetic resonance imaging (MRI). MRI is a non-invasive instrument that is capable of producing high resolution (sub-millimetric) images of the brain using the phenomenon of proton spin resonance. This instrument is sensitive to many physical properties such as spin relaxometry (T1, T2, T2*), spin interactions, spin density or spin displacement. By measuring these properties, a lot of information about the microenvironment can be extracted such as the water/protein/lipid content, the blood flow/volume, the cell density/shape/size, tissue texture, or the content of paramagnetic contrast agent. In these MRI images, GB are clearly shown as heterogeneous tissues (4). The typical imaging features of a GB include an infiltrative, heterogeneous, ring-enhancing lesion with central necrosis and surrounding peritumoral edema. In clinical routine, two main sequences are commonly used to image GB: a contrast-enhanced (CE) T1-weighted image (CE-T1W) that reveals, with gadolinium enhancement, the active regions that present blood-brain barrier breakdown (usually a hyperintense rim with a necrotic core) and a T2-weighted and Fluid Attenuated Inversion Recovery (T2W/FLAIR) image that highlights other tissue abnormalities in the peritumoral region (e.g. tissue softening, water infiltration, vasogenic edema) (4).

Beside anatomic and functional sequences, in vivo proton (^1^H) magnetic-resonance spectroscopic imaging (MRSI), which allows to study metabolic heterogeneity (5), has shown convincing and promising results in GB related to tumor grade and delimitation, prediction of resistance patterns and patient prognosis (6–13). MRSI measures the spatial distribution and concentration of tissue metabolites like choline (Cho) and N-acetyl-aspartate (NAA) which are respectively membrane and neuronal markers, as well as, among others, lactate (necrosis, anaerobic metabolism), creatine/phosphocreatine (ATP metabolism) or myoinositol (putative astroglial marker). An elevated Cho/NAA index (CNI) indicates increased membrane turnover (i.e. cellular proliferation) and reduced neuron density, and is considered to highlight a metabolically active zone of the tumor in high-grade gliomas (14, 15). This metabolic ratio, notably when the CNI is greater than 2 (CNI+), is an interesting predictor of GB patient survival (9–11) and recurrence location (8). Our previous works demonstrated that MRSI is useful in identifying tumor areas with a higher potential for recurrence (6, 7). In our first study (6), we performed the follow-up by MRI and MRSI of 9 patients included in a phase I trial associating tipifarnib and radiotherapy (RT) for GB primary treatment (16) and observed in pre-RT data that CNI+ regions accounted for 25% of CE regions (in CE-T1w sequence) and 10% of regions of T2W/FLAIR hyperintensity corresponding to the infiltrative areas. More interestingly, we showed that the presence of metabolically active CNI+ regions was predictive of the site of relapse after RT, both in CE and FLAIR areas (6). Cho/NAA ratio was also described by other teams as a parameter which may predict earlier GB recurrence (12), shorter progression free survival (PFS) and overall survival (OS) (10), localization of invasive margin of GB (17), but also treatment failure for anti-angiogenic combined therapy (13).

Considering that pre-RT CNI+ GB areas are predictive of the recurrence zones post-treatment (6) and that GIC were demonstrated to mediate recurrence after radio-chemotherapy (18, 19), we hypothesized that these particular metabolic regions (i.e. CNI+) may be enriched in GIC and by this way could favor tumor relapse in these zones. In this regard, recent works have highlighted that higher CNI tumor areas were correlated with stronger Sox2 immunostaining in GB (20). In addition, the MRSI data obtained from tumors generated in mice brain after xenograft of patient-derived GIC unveiled a significant increase in Cho (or Chol-containing compounds) and decrease in NAA (21, 22). Altogether, a noticeable correlation between CNI+ areas and GIC enrichment in GB might be suspected. To answer to this hypothesis, we designed and performed the STEMRI trial (NCT01872221) aiming to study by MRI-MRSI-guided resection the potential enrichment of the CNI+ regions with GIC in comparison with CNI− tumor regions. This trial, whose flow diagram is shown in **Figure 1**, was designed to fully characterize and compare these CNI−/+ regions in both CE and FLAIR zones and then to correlate these data with patients’ clinical outcome after standard radio-chemotherapy treatment.

**Figure 1.**
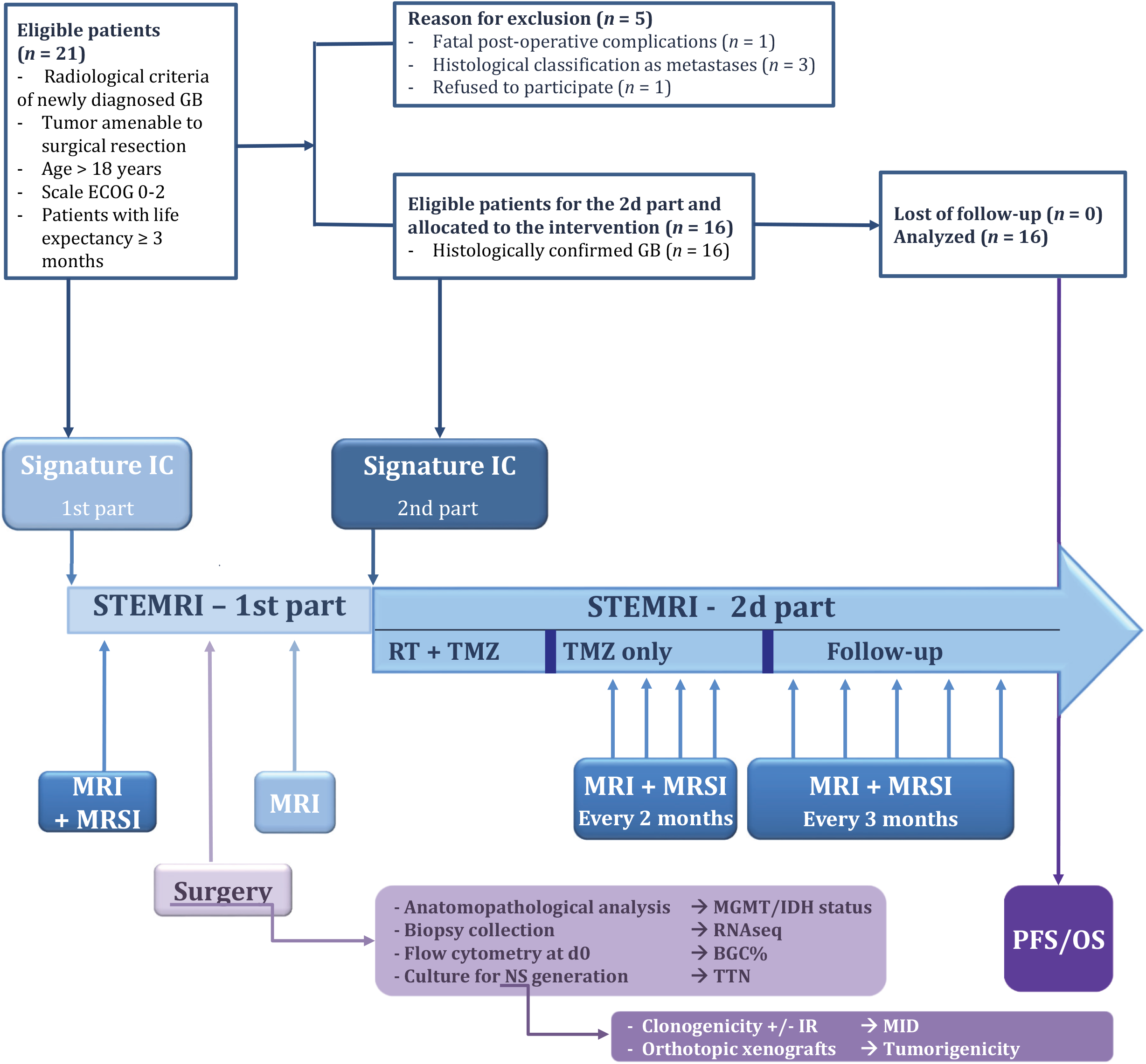
STEMRI trial flow diagram. IC: *Informed Consents*; MRI: *Magnetic Resonance Imaging*; MRSI: *Magnetic Resonance Spectroscopic Imaging*; RT: *Radiotherapy*; TMZ: *Temozolomide*; MGMT: *O*^*6*^*-methylguanine-DNA methyltransferase;* IDH: *Isocitrate Dehydrogenase;* NS: *Neurosphere*; BGC%: *Biopsy GIC Content percentage*; TTN: *Time to Neurosphere formation*; MID: *Mean Inactivation Dose*; PFS: *Progression Free Survival*; OS: *Overall Survival*

## RESULTS

### Patient’s characteristics and clinical outcome

According to inclusion criteria, 21 patients were enrolled in the first part of the study. However, 5 patients could not be included in the second part due to inclusion and exclusion criteria. Three patients were diagnosed after histological analysis as pulmonary, renal and melanoma brain metastases, respectively. One patient experienced postoperative complications that resulted in his death. One patient has decided to withdraw from the trial after surgery in order to receive chemo-radiotherapy sessions in a different hospital center. As a consequence, 16 patients were included in the second part of the trial and evaluable for the entire study. The patient’s characteristics are provided in **Table 1**. Their age ranged between 37 and 78, with a median age of 64 years, 5 females and 11 males. Of note, 43.7% of the patients had a temporal tumor, and percentages for parietal and frontal areas were 18.7% and 37.5%, respectively. All patients had a GB IDH wild type, according to 2021 WHO classification (23). 62.5% were shown to have an unmethylated MGMT (O^6^-methylguanine-DNA methyl-transferase) promoter, 18.7% a methylated MGMT and it was not technically possible to determine MGMT status for 3 patients. All the 16 patients received a cumulative irradiation dose of 60 Gy and concomitant TMZ. At the end of the trial, all the 16 patients experienced GB progression, corresponding to either clinical tumor progression (*n* = 12) or death event (*n* = 4), and all 16 patients had died. The median PFS (progression-free survival) and OS (overall survival) were respectively estimated to 6.5 months [95%CI: 5.5-12.5] and 20.1 months [95%CI: 12.7-25.7] (**Figure 2A**).

**Table 1.** Patients’ characteristics. *nd*: not determined.

| Patient number | Tumor name | Sex | Age Range (years) | Tumor localization | Treatment |  |  | Nature of the progression | Patient state | Progression-free survival (months) | Overall survival (months) | MGMT methylation status (Y/N) | IDH1 status |
| --- | --- | --- | --- | --- | --- | --- | --- | --- | --- | --- | --- | --- | --- |
|  |  |  |  |  | Irradiation dose (Gy) | Concomittant TMZ dose (mg/m²) | Adjuvant TMZ cycles |  |  |  |  |  |  |
| sub-001 | SRA | M | 76-80 | temporal | 60 | 3150 | 2 | PROGRESSION | Deceased | 6.21 | 8.05 | N | Wt |
| sub-002 | SRB | F | 76-80 | parietal | 60 | 3150 | 0 | PROGRESSION | Deceased | 3.68 | 21.32 | N | Wt |
| sub-003 | SRC | M | 66-70 | temporal | 60 | 3150 | 6 | PROGRESSION | Deceased | 14.98 | 30.65 | Y | Wt |
| sub-006 | SRF | F | 71-75 | temporal | 60 | 1950 | 0 | DEATH (heart attack) | Deceased | 2.76 | 2.76 | N | Wt |
| sub-007 | SRG | M | 56-60 | frontal | 60 | 3300 | 2 | PROGRESSION | Deceased | 6.11 | 16.95 | Y | Wt |
| sub-008 | SRH | F | 56-60 | parietal | 60 | 3150 | 6 | PROGRESSION | Deceased | 21.16 | 36.6 | Y | Wt |
| sub-009 | SRI | F | 71-75 | frontal | 60 | 3525 | 6 | PROGRESSION | Deceased | 12.45 | 29.34 | *nd* | Wt |
| sub-010 | SRJ | M | 51-55 | frontal | 60 | 3150 | 4 | DEATH | Deceased | 15.11 | 20.4 | N | Wt |
| sub-012 | SRK | M | 61-65 | temporal | 60 | 3150 | 5 | PROGRESSION | Deceased | 9.1 | 27.27 | N | Wt |
| sub-013 | SRL | F | 56-60 | temporal | 60 | 3225 | 1 | DEATH | Deceased | 5.19 | 6.87 | N | Wt |
| sub-014 | SRM | M | 71-75 | temporal | 60 | 3150 | 3 | PROGRESSION | Deceased | 5.45 | 18.56 | *nd* | Wt |
| sub-015 | SRN | M | 71-75 | temporal | 60 | 3150 | 3 | PROGRESSION | Deceased | 5.55 | 20.11 | N | Wt |
| sub-017 | SRO | M | 46-50 | frontal | 60 | 2775 | 6 | PROGRESSION | Deceased | 10.74 | 25.69 | N | Wt |
| sub-019 | SRQ | M | 76-80 | frontal | 60 | 2775 | 0 | DEATH (nocardiosis) | Deceased | 21.09 | 21.09 | *nd* | Wt |
| sub-020 | SRR | M | 36-40 | frontal | 60 | 3525 | 3 | PROGRESSION | Deceased | 6.47 | 14.26 | N | Wt |
| sub-021 | SRS | M | 56-60 | parietal | 60 | 3150 | 5 | PROGRESSION | Deceased | 8.67 | 12.68 | N | Wt |

**Figure 2.**
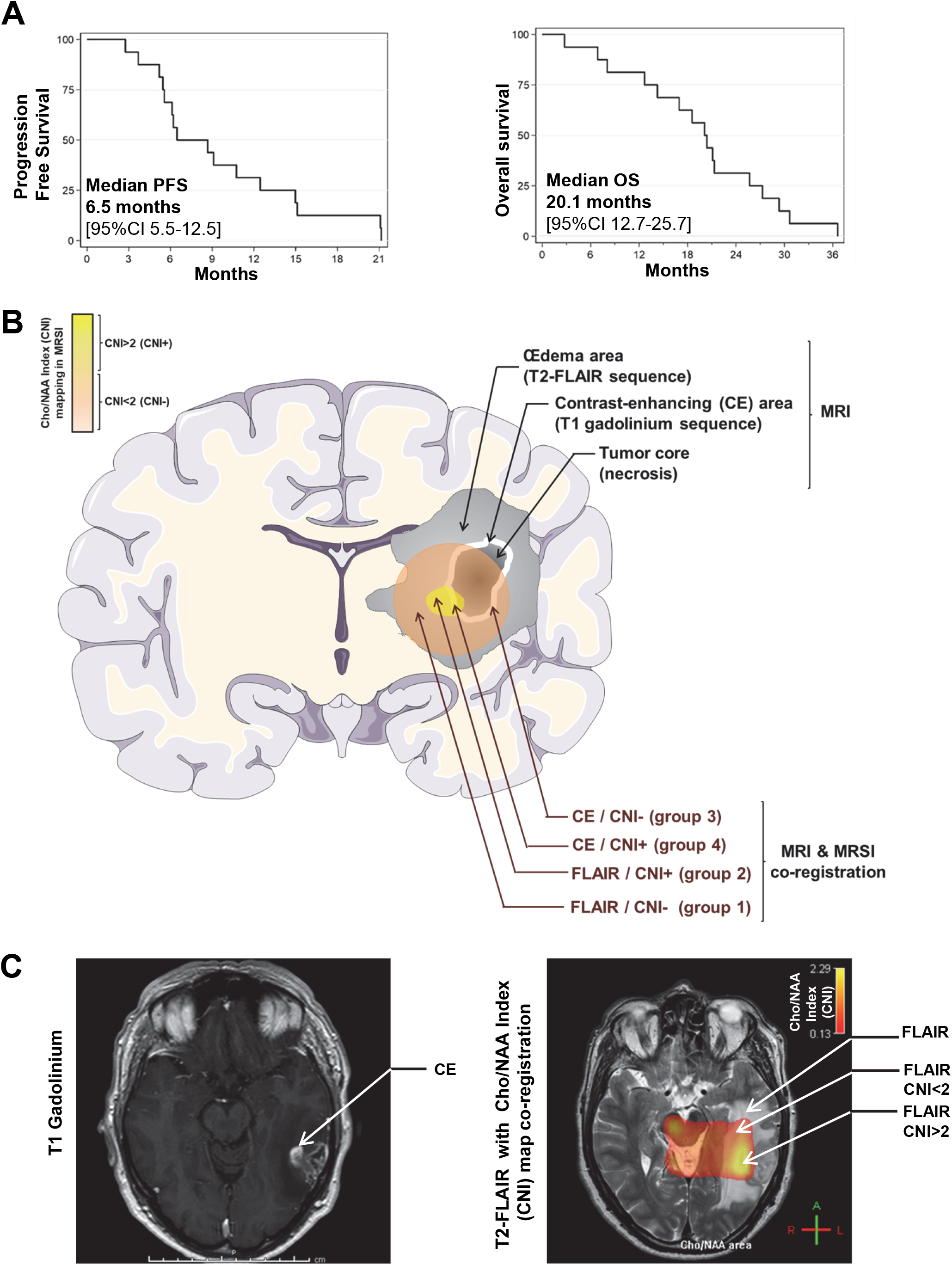
STEMRI patients’ survival analyses and related study design. **(A)** Kaplan-Meier curves show Progression Free Survival (PFS, left panel) and Overall Survival (OS, right panel) of GB patients enrolled in the SEMRI trial (n = 16). **(B-C)** General descriptive scheme **(B)** and MRI/MRSI representative (sub-003 patient) images **(C)** of the study strategy for surgical samples collection. Pre-operative MRI/MRSI co-registration allowed the determination of two CNI (Choline/N-Acetyl-Aspartate Index) metabolic zones (CNI<2 and CNI>2) in both CE (Contrast Enhancement) and FLAIR (Fluid Attenuated Inversion Recovery) areas, determined through T1 gadolinium and T2-FLAIR MRI sequences, respectively. A maximum of four different biopsies were then collected for each patient in the above described zones: FLAIR/CNI− (group 1); FLAIR/CNI+ (group 2); CE/CNI− (group 3); CE/CNI+ (group 4).

### Patients Biopsies

All the 16 patients had different tumor biopsies performed according to the study for analysis and for generation of derived GIC-enriched neurospheres. For each patient, 4 samples, when possible, were biopsied in 4 different tumor areas: FLAIR/CNI− (group 1); FLAIR/CNI+ (group 2); CE/CNI− (group 3); CE/CNI+ (group 4), as described in **Figure 2B-C** and in Methods. The details of the biopsies per patients are shown in **Table 2**. To summarize, 54 biopsies samples were collected for 16 patients. These biopsies samples were divided immediately after surgery in three parts for (i) RNA extraction (51/54 biopsies generated optimal quality RNA for RNASeq), (ii) FACS analysis of GIC subpopulation at day 0 (54/54), and (iii) long term culture in stem cell medium to generate NS. In these restrictive culture conditions, 42/54 processed biopsies led to stable primary NS generation and, among them, 34/54 were able to generate secondary NS in culture (**Table 2**).

**Table 2.** Tumor samples’ characteristics. *nd*: not determined.

| Patient number | Tumor Name | Tumor Localization | Biopsies Y/N | Sample Name | Biopsy GIC Content (BGC) at day 0 (%) | Biopsy RNA Y/N | Biopsy Molecular Classification |  | Stable Primary Neurosphere generation Y/N | Time to Neurosphere (TTN) generation (days) | Stable Secondary Neurosphere generation Y/N |
| --- | --- | --- | --- | --- | --- | --- | --- | --- | --- | --- | --- |
|  |  |  |  |  |  |  | Verhaak 2010 | Wang 2017 |  |  |  |
|  |  |  |  |  |  |  | PN: proneural<br>Ne: neural<br>Mes: mesenchymal<br>CL: classical | PN: proneural<br>Mes: mesenchymal<br>CL: classical |  |  |  |
| sub-001 | SRA | (1) FLAIR/CNI- | N | - | - | - | - | - | - | - | - |
|  |  | (2) FLAIR/CNI+ | Y | SRA4 | 6.08 | Y | PN | PN | Y | 13 | Y |
|  |  | (3) CE/CNI- | Y | SRA5 | 7.02 | Y | PN | PN | Y | 8 | Y |
|  |  | (4) CE/CNI+ | N | - | - | - | - | - | - | - | - |
| sub-002 | SRB | (1) FLAIR/CNI- | Y | SRB3 | 10.56 | Y | Ne | PN | Y | 36 | Y |
|  |  | (2) FLAIR/CNI+ | N | - | - | - | - | - | - | - | - |
|  |  | (3) CE/CNI- | Y | SRB1 | 20.1 | Y | Mes | Mes | Y | 50 | Y |
|  |  | (4) CE/CNI+ | Y | SRB2 | 44.57 | Y | Mes | Mes | Y | 11 | Y |
| sub-003 | SRC | (1) FLAIR/CNI- | Y | SRC1 | 6 | Y | Ne | PN | Y | 46 | Y |
|  |  | (2) FLAIR/CNI+ | Y | SRC2 | 12.1 | Y | Ne | PN | Y | 16 | Y |
|  |  | (3) CE/CNI- | Y | SRC3 | 40.15 | Y | Ne | CL | Y | 16 | Y |
|  |  | (4) CE/CNI+ | Y | SRC4 | 48.8 | Y | CL | PN | Y | 53 | Y |
| sub-006 | SRF | (1) FLAIR/CNI- | Y | SRF1 | 9.03 | Y | Ne | CL | N | - | N |
|  |  | (2) FLAIR/CNI+ | Y | SRF5 | 22.97 | Y | CL | CL | Y | 11 | N |
|  |  | (3) CE/CNI- | Y | SRF3 | 4.78 | Y | Mes | Mes | N | - | N |
|  |  | (4) CE/CNI+ | Y | SRF4 | 9.22 | Y | PN | Mes | N | - | N |
| sub-007 | SRG | (1) FLAIR/CNI- | Y | SRG5 | 9.1 | Y | Ne | PN | Y | 60 | N |
|  |  | (2) FLAIR/CNI+ | Y | SRG4 | 10.8 | Y | Ne | Mes | Y | 21 | Y |
|  |  | (3) CE/CNI- | Y | SRG1 | 39 | Y | Mes | Mes | Y | 2 | Y |
|  |  | (4) CE/CNI+ | Y | SRG2 | 22.7 | Y | Mes | Mes | Y | 2 | Y |
| sub-008 | SRH | (1) FLAIR/CNI- | Y | SRH1 | 4.39 | Y | Mes | CL | *nd* | *nd* | N |
|  |  | (2) FLAIR/CNI+ | Y | SRH3 | 2.52 | Y | CL | CL | Y | 19 | Y |
|  |  | (3) CE/CNI- | Y | SRH2 | 16.49 | Y | CL | CL | Y | 5 | Y |
|  |  | (4) CE/CNI+ | Y | SRH4 | 10.18 | N | *nd* | *nd* | N | - | N |
| sub-009 | SRI | (1) FLAIR/CNI- | N | - | - | - | - | - | - | - | - |
|  |  | (2) FLAIR/CNI+ | N | - | - | - | - | - | - | - | - |
|  |  | (3) CE/CNI- | N | - | - | - | - | - | - | - | - |
|  |  | (4) CE/CNI+ | Y | SRI4 | 13.58 | Y | CL | Mes | Y | 1 | Y |
| sub-010 | SRJ | (1) FLAIR/CNI- | Y | SRJ1 | 5.91 | Y | Ne | PN | N | - | N |
|  |  | (2) FLAIR/CNI+ | Y | SRJ2 | 10.02 | Y | PN | PN | N | - | N |
|  |  | (3) CE/CNI- | Y | SRJ3 | 16.42 | Y | Mes | Mes | N | - | N |
|  |  | (4) CE/CNI+ | Y | SRJ4 | 54.13 | Y | Mes | Mes | Y | 5 | N |
| sub-012 | SRK | (1) FLAIR/CNI- | Y | SRK3 | 6.02 | Y | Ne | PN | N | - | N |
|  |  | (2) FLAIR/CNI+ | Y | SRK4 | 11.67 | N | *nd* | *nd* | N | - | N |
|  |  | (3) CE/CNI- | Y | SRK1 | 60.26 | Y | Mes | Mes | Y | 8 | Y |
|  |  | (4) CE/CNI+ | Y | SRK2 | 68.53 | Y | CL | CL | Y | 8 | Y |
| sub-013 | SRL | (1) FLAIR/CNI- | Y | SRL4 | 23.27 | Y | Ne | PN | Y | 21 | Y |
|  |  | (2) FLAIR/CNI+ | Y | SRL3 | 36.27 | Y | Ne | PN | Y | 13 | Y |
|  |  | (3) CE/CNI- | Y | SRL1 | 63.64 | Y | Mes | CL | Y | 13 | Y |
|  |  | (4) CE/CNI+ | Y | SRL2 | 79.55 | Y | Ne | CL | Y | 21 | Y |
| sub-014 | SRM | (1) FLAIR/CNI- | Y | SRM1 | 14.02 | N | *nd* | *nd* | Y | 18 | Y |
|  |  | (2) FLAIR/CNI+ | Y | SRM3 | 16.03 | Y | Ne | PN | Y | 19 | Y |
|  |  | (3) CE/CNI- | N | - | - | - | - | - | - | - | - |
|  |  | (4) CE/CNI+ | Y | SRM2 | 42.07 | Y | CL | CL | Y | 5 | Y |
| sub-015 | SRN | (1) FLAIR/CNI- | Y | SRN3 | 3.87 | Y | Ne | PN | N | - | N |
|  |  | (2) FLAIR/CNI+ | Y | SRN4 | 15.94 | Y | PN | PN | Y | 4 | Y |
|  |  | (3) CE/CNI- | Y | SRN1 | 3.97 | Y | PN | PN | Y | 11 | Y |
|  |  | (4) CE/CNI+ | Y | SRN2 | 13.04 | Y | PN | PN | Y | 28 | Y |
| sub-017 | SRO | (1) FLAIR/CNI- | Y | SRO3 | 10.71 | Y | Ne | CL | Y | 14 | Y |
|  |  | (2) FLAIR/CNI+ | Y | SRO2 | 5.45 | Y | PN | PN | Y | 10 | Y |
|  |  | (3) CE/CNI- | Y | SRO1 | 7.46 | Y | Mes | Mes | Y | 5 | Y |
|  |  | (4) CE/CNI+ | N | - | - | - | - | - | - | - | - |
| sub-019 | SRQ | (1) FLAIR/CNI- | Y | SRQ2 | 5.55 | Y | Ne | PN | Y | 33 | N |
|  |  | (2) FLAIR/CNI+ | N | - | - | - | - | - | - | - | - |
|  |  | (3) CE/CNI- | Y | SRQ3 | 42.05 | Y | PN | PN | Y | 33 | N |
|  |  | (4) CE/CNI+ | N | - | - | - | - | - | - | - | - |
| sub-020 | SRR | (1) FLAIR/CNI- | Y | SRR2 | 4.04 | Y | CL | CL | Y | 13 | N |
|  |  | (2) FLAIR/CNI+ | Y | SRR4 | 19.11 | Y | CL | CL | Y | 13 | N |
|  |  | (3) CE/CNI- | Y | SRR3 | 4.92 | Y | Ne | PN | Y | 18 | N |
|  |  | (4) CE/CNI+ | Y | SRR1 | 10.69 | Y | CL | CL | Y | 15 | N |
| sub-021 | SRS | (1) FLAIR/CNI- | Y | SRS3 | 4.7 | Y | Ne | PN | Y | 8 | Y |
|  |  | (2) FLAIR/CNI+ | Y | SRS2 | 13.37 | Y | CL | CL | Y | 8 | Y |
|  |  | (3) CE/CNI- | Y | SRS1 | 12.87 | Y | CL | CL | Y | 8 | Y |
|  |  | (4) CE/CNI+ | Y | SRS4 | 8.34 | Y | PN | PN | Y | 8 | Y |

### GIC quantification in patient’s biopsies

We compared by FACS the percentages of the GIC subpopulation in each biopsy just after surgery (day 0) between FLAIR/CNI− and FLAIR/CNI+ as well as between CE/CNI− and CE/CNI+ tumor areas. For the 12 pairs of biopsies CNI−/CNI+ in the FLAIR area, the Biopsy GIC content at D0 (BGC%) was significantly higher in the CNI+ biopsies (**Figure 3A and supplemental Table 1)**. However no significant difference could be observed in the CE zone between CNI− and CNI+ biopsies (**Figure 3B and supplemental Table 1**). Beyond pairs, when all the samples were considered, we observed similar results between CNI− and CNI+ in FLAIR and CE areas, respectively (**supplemental Figure 1A**). Of note, when all biopsies of the CNI− and CNI+ were considered, regardless of their FLAIR or CE origin, the observed increase of BGC% in CNI+ appeared to be not significant. On the contrary, if we compare CE (tumoral) and FLAIR (peritumoral) zones, regardless of their CNI status, a strong increase of the BGC% in CE biopsies can be highlighted (**supplemental Figure 1A**). We also analyzed by FACS the expression of stem and differentiation markers in the GIC population in each biopsy. Only a non-significant increase in the stem markers CD44, CD15 and Nestin could be observed between CNI− and CNI+ in FLAIR, but no clear tendency between CNI− and CNI+ in CE zone (**Figure 3C-D**) or regardless of the FLAIR or CE origin (**supplemental Figure 1B**). However, CE biopsies, regardless of their CNI status, comprise GIC overexpressing stem markers Nestin, CD44, Sox2, ITGB8 (3, 24), in comparison with GIC contained in FLAIR biopsies, which seem to overexpress the oligodendrocytic marker O4 (**supplemental Figure 1B**).

**Figure 3.**
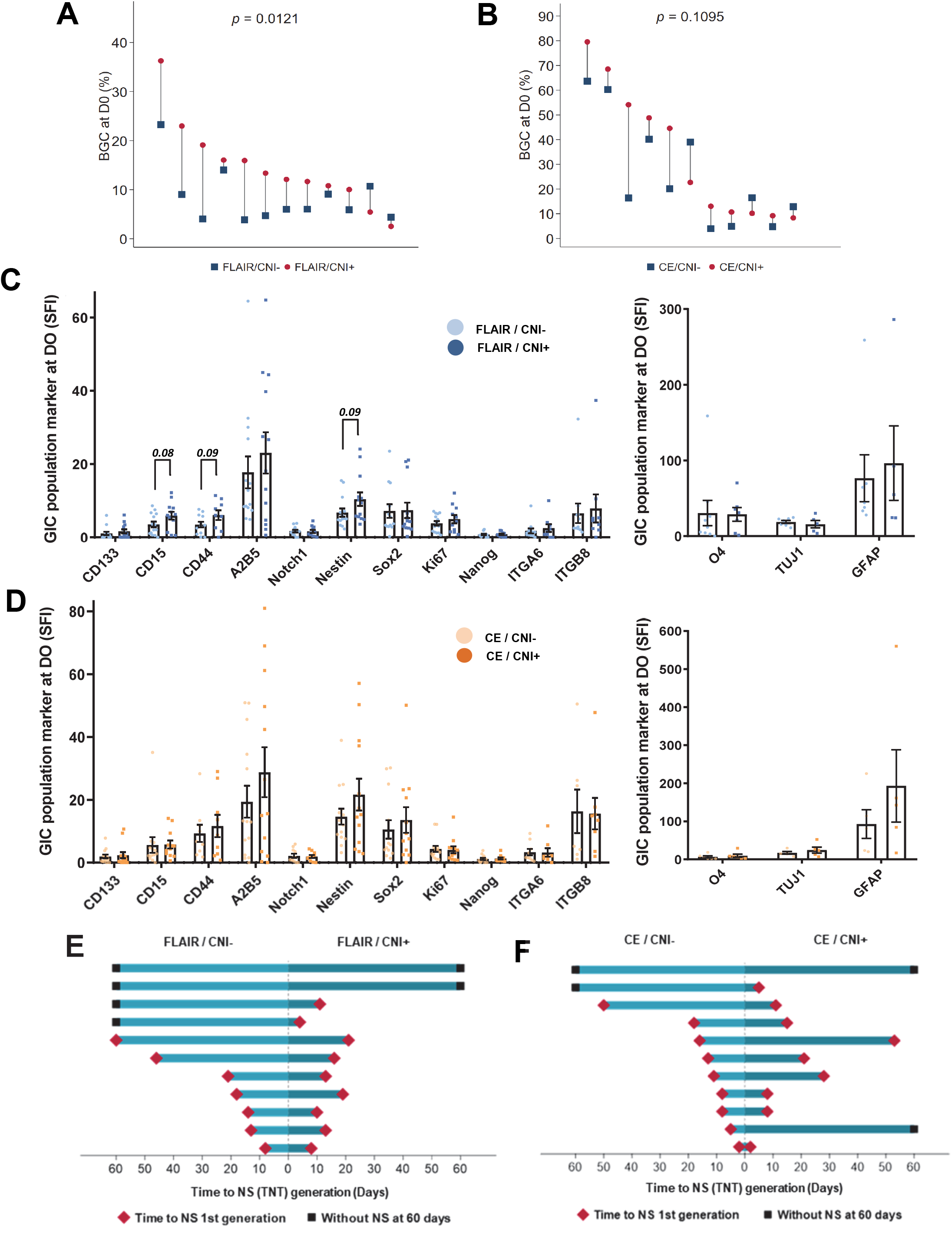
FLAIR/CNI+ biopsies are characterized by an enrichment in the GIC subpopulation and a shorter time to neurosphere (NS) generation compared to FLAIR/CNI− biopsies. **(A-B)** Pairwise comparison of the Biopsy GIC Content at day 0 (D0) after surgery (BGC%), as determined by FACS, either in FLAIR/CNI− and FLAIR/CNI+ **(A)** or in CE/CNI− and CE/CNI+ **(B)** samples. Detailed pair by pair comparisons between the two biopsy groups, associated to statistical analysis, were shown. **(C-D)** Immunofluorescence analysis performed by FACS for the indicated stem (left panels) and differentiation (right panels) markers in the GIC subpopulation analyzed at D0 either in FLAIR/CNI− and FLAIR/CNI+ **(C)** or in CE/CNI− and CE/CNI+ **(D)** samples. The SFI (Specific Fluorescence Index), used to evaluate the marker expression level, were expressed as means ± SEM of all analyzed samples. **(E-F)** Pairwise comparison of the Time to NS (TTN) generation, either in FLAIR/CNI− and FLAIR/CNI+ **(E)** or in CE/CNI− and CE/CNI+ **(F)** samples.

### NS generation from patient’s biopsies

According to the FACS analysis of GIC content in biopsies, there is a notable intra-patient heterogeneity in FLAIR tumor areas between CNI− and CNI+ zones. In this inflammatory and edematous peritumor area, CNI+ biopsies appeared to be enriched in GIC compared to CNI− biopsies. As a consequence, we also studied whether or not these biopsies gave rise to GIC-enriched NS more or less rapidly depending on their CNI origin. By determining the Time to NS (TTN) formation (in days), we noticed, in our 11 analyzable pairs of FLAIR biopsies, a clear tendency for FLAIR/CNI+ samples to generate NS faster (Median = 13 days) compared to their CNI− counterparts (18 days) (**Figure 3E and supplemental Table 2**). Beyond paired biopsies, when all the samples are considered, a significant shortening of the TTN was observed in FLAIR/CNI+ compared to FLAIR/CNI− biopsies, very closed to the TTN observed in CE/CNI− and CE/CNI+ groups (**supplemental Table 2 and supplemental Figure 2**). Of note, between these two last groups, no significant differences could be showed (**Figure 3F, supplemental Table 2 and supplemental Figure 2**), as well as between CNI− and CNI+ groups (regardless of their CE/FLAIR origin) and between FLAIR and CE groups (regardless of their CNI status) (**supplemental Figure 2**). In line with a faster appearance of GIC-enriched NS in FLAIR/CNI+ samples, we found that FLAIR/CNI− biopsies could generate stable primary and secondary NS at 69.2% and 42.9%, respectively, while FLAIR/CNI+ biopsies showed a better yield, with 84.6% and 69.2%, respectively. Of note, the NS generation percentages in FLAIR/CNI+ samples are quite similar with those observed in CE biopsies, regardless of their CNI status (**supplemental Table 2**). So, concerning the pattern of NS generation, it appears that FLAIR/CNI+ tumor zones, contrary to FLAIR/CNI−, may contain a sufficient subpopulation of GIC to produce GIC-enriched NS, similarly to CE biopsies, which contain a greater subpopulation of GIC.

### Tumorigenicity potential of GIC coming from different areas

We have also studied the tumorigenic potential of the secondary NS generated from each tumor areas, since GB cells tumorigenicity in orthotopically-xenografted nude mice was shown as a major property of GIC (3). For each primary GIC lines able to generate stable secondary NS (n = 34/54), NS-dissociated cells were subjected to orthotopic xenografts in nude mice (n = 4-6 mice per NS samples) in order to assess their tumorigenic potential. They all generated brain tumors to varying degrees and these GIC-derived tumors were analyzed by hemalun staining and Nestin immunostaining to confirm their tumorigenic ability (**supplemental Figure 3**). By analyzing the global median OS in xenografted mice as a quantitative variable (in days [95% CI]), we observed that GIC obtained from FLAIR/CNI− NS were associated with an increased median OS (206 [127;420]), compared to the mice implanted with FLAIR/CNI+ NS-derived cells (182 [145;232]) or with CE NS-derived cells (189 [142;227] for CE/CNI− and 188 [152;270] for CE/CNI+) (**supplemental Figure 4**). Univariable analysis by Logrank test confirmed that OS in mice xenografted with FLAIR/CNI+ NS-derived cells is significantly lower than in the FLAIR/CNI− group (**Figure 4B**), with a survival rate at t=250 days of 42.9% *vs* 17.5% for FLAIR/CNI− and FLAIR/CNI+ groups, respectively (**supplemental Table 3**). On the contrary, no significant differences could be highlighted between CE/CNI− and CE/CNI+ groups, between CNI− and CNI+ mice (irrespective of the FLAIR/CE origin) or between CE and FLAIR conditions (irrespective of the CNI pattern) (**Figure 4C-E**). Then, FLAIR/CNI+ derived NS, which are generated *in vitro* from tumor biopsies faster than FLAIR/CNI− samples (13.36 +/− 1.54 *vs* 27.67 +/− 5.78 as day averages +/− SEM, respectively), also appear to give rise to tumors more rapidly in xenografted mice.

**Figure 4.**
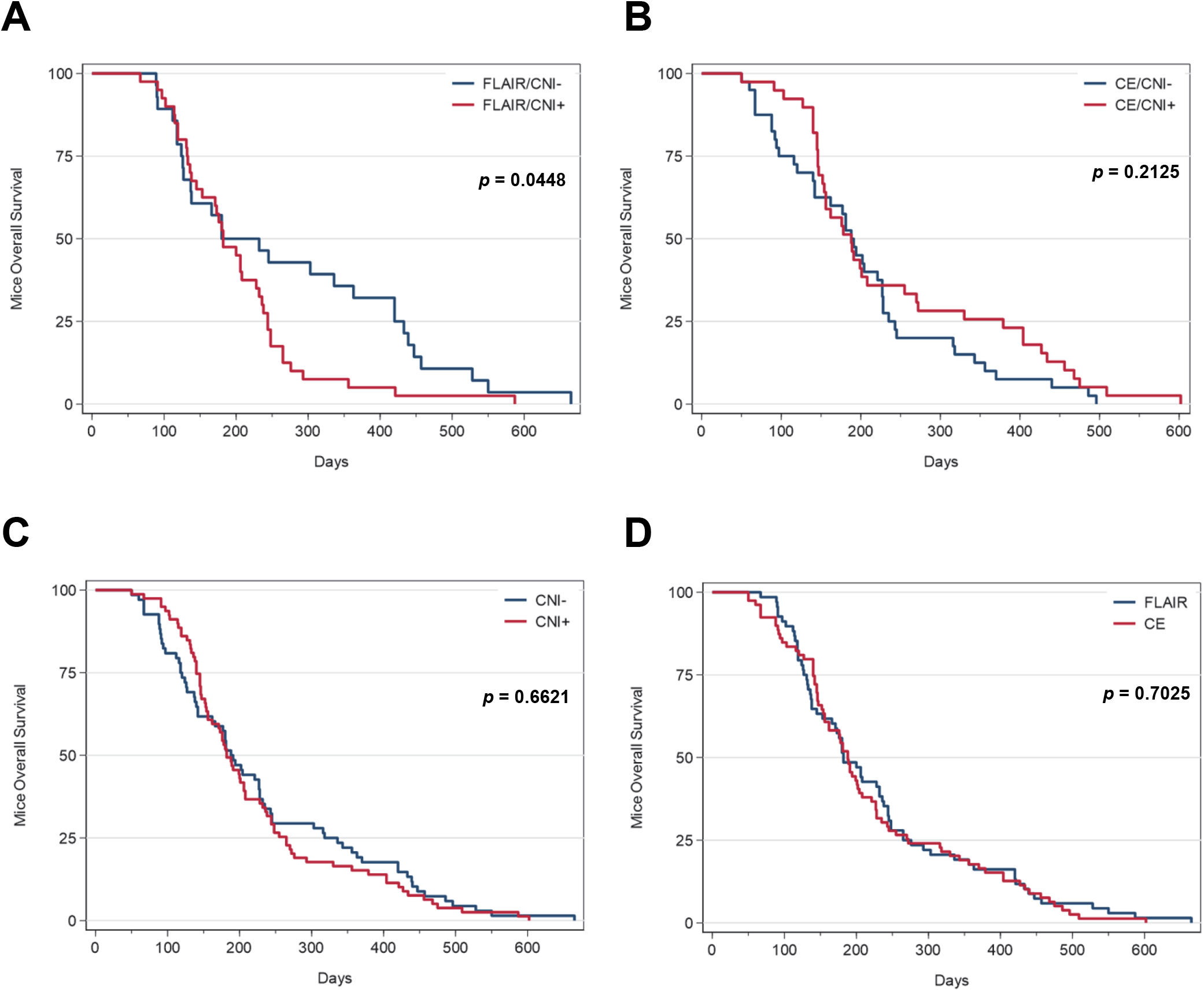
GIC primary cell lines obtained from FLAIR/CNI+ biopsies are associated with increased tumorigenesis in xenografted mice compared to FLAIR/CNI− neurospheres. **(A-E)** For each primary GIC cell lines able to generate stable secondary neurospheres, NS-dissociated cells were subjected to orthotopic xenografts in nude mice (n = 4-6 mice per sample). Overall survival (OS) data were determined for each GIC groups (FLAIR/CNI−; FLAIR/CNI+; CE/CNI−; CE/CNI+) in order to assess their tumorigenic ability. **(A-D)** Kaplan-Meier curves for OS in mice xenografted with the indicated GIC subgroups. Exact p-values for FLAIR/CNI− vs FLAIR/CNI+ **(A)**, CE/CNI− vs CE/CNI+ **(B)**, CNI− vs CNI+ **(C)** or FLAIR vs CNI **(D)** group comparisons were determined by logrank analysis.

### Radiosensitivity of GIC according to different zones of biopsy

As GIC phenotype is associated with increased radioresistance and since we have shown that CNI2 areas were predictive of the site of relapse after radiotherapy, we also checked whether GIC cell lines coming from CNI+ area had a decrease in radiosensitivity compared to those coming from CNI− area.

For that, the *in vitro* radiosensitivity of each stable NS cell line was studied by realizing clonogenic assays at increasing irradiation doses and subsequently establishing their MID (mean inactivation dose). We did not observe any significant differences between FLAIR/CNI− and FLAIR/CNI+ NS or between CE/CNI− and CE/CNI+ NS **(supplemental Table 4)**.

### Relationship between GIC-associated pattern in CNI−/+ biopsies and patient outcome

FLAIR/CNI+ tumor areas, which were found by our team to predict the site of GB relapse after chemo/radiotherapy, appeared to be enriched in GIC and to give rise more rapidly to stable NS in restrictive culture conditions, compared to FLAIR/CNI− biopsies. In order to find out a possible link between the previously described GIC-related parameters in the biopsies and patient outcome, we studied OS and PFS for our 16 patients according to their Biopsy GIC Content (BGC%, determined by FACS at day 0) or to their Time to NS (TTN) generation. Concerning BGC and TTN, patients were dichotomized into two distinct groups according to the median values observed in FLAIR/CNI+ biopsies. Univariable analysis of OS and PFS by Logrank test highlighted a significant association of high BGC with decreased OS and PFS (**Figure 5A-B and supplemental Table 5A-B**). This was confirmed by analysis through Cox statistical model using continuous variables (**supplemental Table 5A-B**). Of note, if we use for each patient the maximum BGC content of both FLAIR biopsies (CNI− and CNI+), the same type of association between low OS/PFS and high BGC can be described with the Cox model and the Logrank test, with a loss of statistical significance in the latter case (**supplemental Table 5A-B**). Focusing on the TTN parameter, univariable analyses using Cox model did not show a significant association between OS/PFS and TTN in days, although there is a trend in this direction for FLAIR/CNI+ samples (**supplemental Table 6A-B)**. We then refined this TTN parameter to analyze both OS and PFS according to their ability to generate or not stable Primary NS at 14 days after surgery and culture in stem medium (PN14), since only 27.3% of FLAIR/CNI− biopsies could generate NS at 14 days compared to 50% in FLAIR/CNI+ (**supplemental Table 2**). Logrank test analysis of OS/PFS according to PN14 parameter in FLAIR/CNI+ depicted a significant association of PN14+ capacity with a decreased OS (*p = 0*.*0190*) and a strong tendency with a reduced PFS (*p = 0*.*0606*) (**Figure 5C-D and supplemental Table 7A-B**). Of note, if FLAIR/CNI+ GIC-related parameters (BGC and PN14) were correlated with reduced OS and PFS, it was not the case for CE biopsies, regardless of their CNI status (**supplemental Tables 5 and 7**). Consequently, FLAIR/CNI+ tumor areas specifically appear to be predictive of GB patient outcome when considering their relative enrichment in GIC.

**Figure 5.**
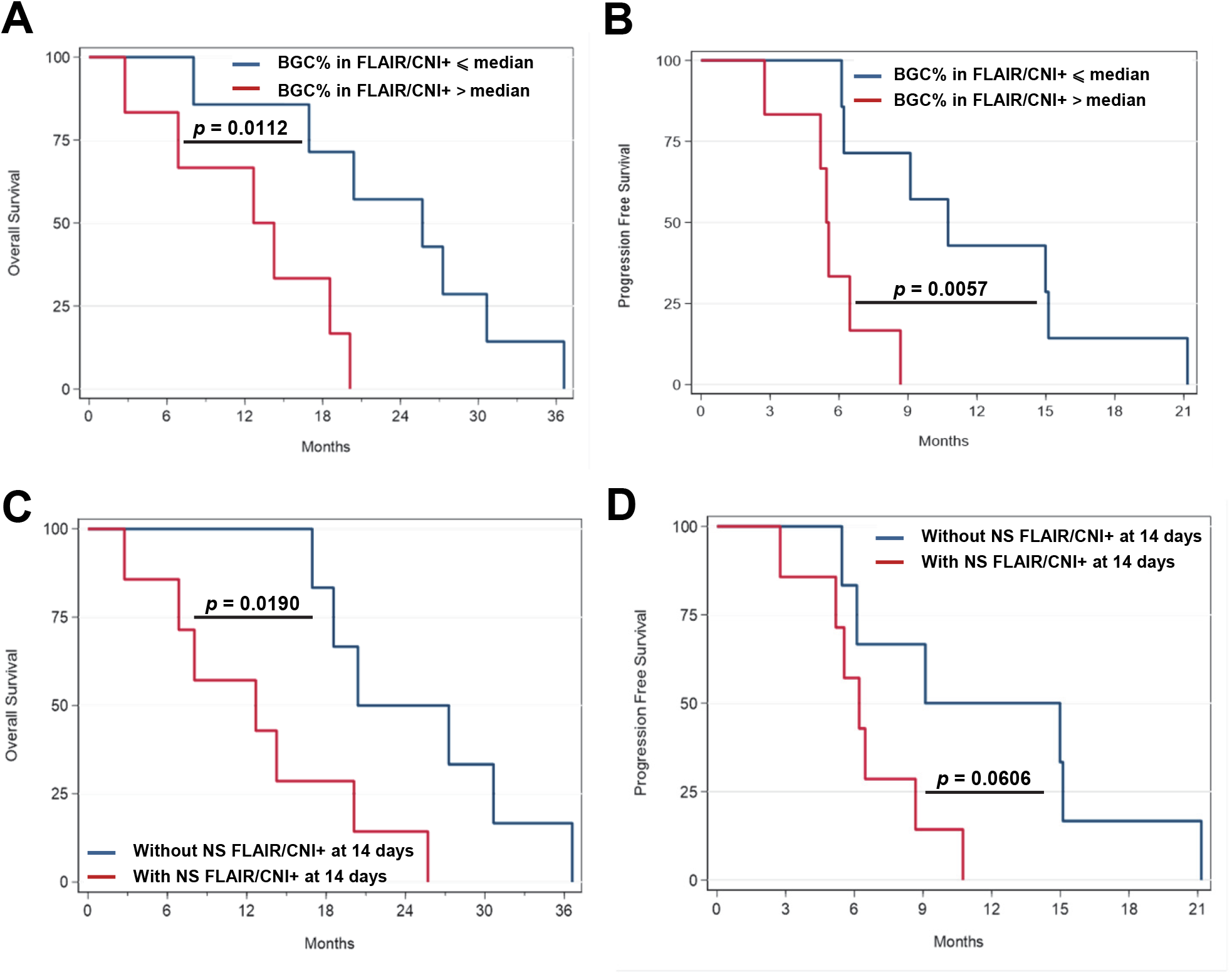
Higher Biopsy GIC content (BGC) and increased ability to promptly generate primary neurospheres in FLAIR/CNI+ samples are two biological parameters associated with worse patient outcome. **(A-B)** Kaplan-Meier curves for OS **(A)** and PFS **(B)** in GB patients enrolled in the SEMRI trial when dichotomized into two distinct groups according to the median BGC values observed in FLAIR/CNI+ biopsies. **(C-D)** Kaplan-Meier curves for OS **(C)** and PFS **(D)** in GB patients when dichotomized into two distinct groups according to FLAIR/CNI+ biopsies’ ability to generate or not stable primary NS in stem medium at 14 days after surgery. **(A-D)** Exact p-values were determined by logrank analysis.

### RNASeq analysis of patient’s biopsies

In light of the previous results, it appears that FLAIR/CNI+ tumor areas, predictive of the site of relapse in GB patients, possess specific biological (higher GIC content, faster primary NS generation), preclinical (increased tumorigenicity in mice) and clinical properties (correlation of patients PFS/OS with biological parameters). In order to observe these clinico-biological results through the prism of molecular characterization, we performed a RNA-sequencing analysis of all available biopsies (51/54 samples), including 13/14 FLAIR/CNI−, 12/13 FLAIR/CNI+, 14/14 CE/CNI− and 12/13 CE/CNI+ samples. Expression dataset were first used to determine the molecular subtype of each biopsy according to either Verhaak or Wang classification (25, 26). Using Verhaak classification, samples could be classified into 4 molecular subgroups (Ne: Neural; Pn: proneural; Mes: Mesenchymal; Cl: Classical) and the results show that CE biopsies are mainly from Mes and Cl subclasses (62.96%) and quite homogenous between CNI− and CNI+ areas (**Figure 6A**). However, in FLAIR peritumor zone, we observe a majority of Pn and Ne subclasses (70.37%) and a marked difference between FLAIR/CNI− and FLAIR/CNI+ zones. Indeed, FLAIR/CNI− biopsies were predominantly classified as Ne subtype (78.57%), while FLAIR/CNI+ tumor samples were divided into three equivalent groups (30.77% of Pn, Ne and Cl subtypes). As demonstrated by Wang et al in 2017, it appeared that the Neural subgroup is in fact non-tumor specific and is due to normal neural lineage contamination, frequently encountered at the tumor margin (26). In light of this observation, it is worthwhile to consider that Ne subclass is overrepresented in FLAIR/CNI− biopsies compared to FLAIR/CNI+ samples, which could mean that FLAIR/CNI+ peritumor areas are enriched in cancer cells contrary to FLAIR/CNI− zones. We then employed the Wang Classification, using the Gliovis data portal (27), to refine the molecular classification into 3 different and more tumor-specific subgroups: Pn, Mes and Cl. As shown in Fig. 5A, FLAIR biopsies were mainly classified as Pn (59.26%) and Cl (29.63%), and CE samples as Mes (40.74%), Cl (29.63%) and Pn (25.93%). It should be noted that no significant differences in the molecular Wang subclasses could be observed when comparing CNI− and CNI+ biopsies, either in FLAIR, CE or FLAIR+CE areas (**Figure 6A**).

**Figure 6 (part 1/2).**
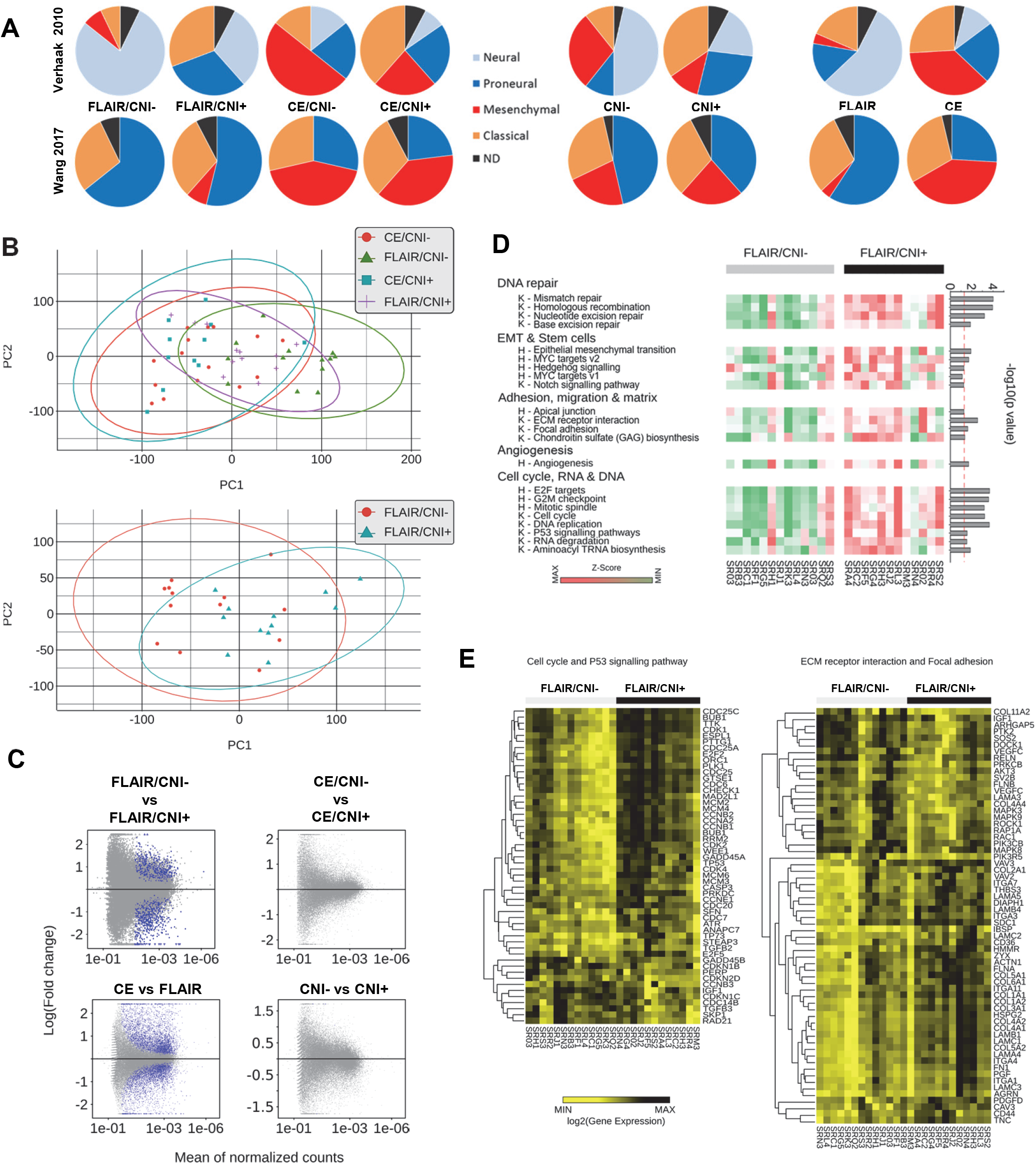
RNAseq analyses revealed that FLAIR/CNI+ biopsies were enriched in gene signatures related to stemness/EMT, migration/adhesion and cell cycle/DNA repair pathways compared to FLAIR/CNI+ tumor samples. RNAseq analyses were conducted on the 51 available tumor biopsies obtained from 16 GB patients and segregated into 4 different groups (FLAIR/CNI−; FLAIR/CNI+; CE/CNI−; CE/CNI+). **(A)** Molecular classification of GB biopsies of each indicated group according to either Verhaak (upper diagrams) or Wang (lower diagrams) classification. ND: not determined. **(B)** Principal Component Analysis (PCA) of RNAseq profiles of the 4 different biopsy groups (upper panel) or restricted to FLAIR/CNI− and FLAIR/CNI+ samples only (lower panel). **(C)** MA plots were generated after a differential expression analysis between the indicated biopsy groups, based on the thresholds of an adjusted q-value < 0.05 and a log2 fold-change ≥ 2. Blue dots indicate differentially expressed genes. **(D-E)** KEGG (K) and HALLMARK (H) pathway enrichment analyses were performed on FLAIR/CNI− vs FLAIR/CNI+ biopsies and showed the statistically significant upregulation of indicated pathways **(D)**. Differentially expressed genes in specified KEGG pathways were also represented in a supervised analysis between these two groups **(E). (F-G)** Gene Ontology (GO) Biological Processes **(F)** and REACTOME **(G)** pathway enrichment analyses were performed on FLAIR/CNI− vs FLAIR/CNI+ biopsies and showed the statistically significant differential expression pattern of specified gene signatures through both supervised (left panel) and unsupervised (right panel) analyses.

**Figure 6 (part 2/2).**
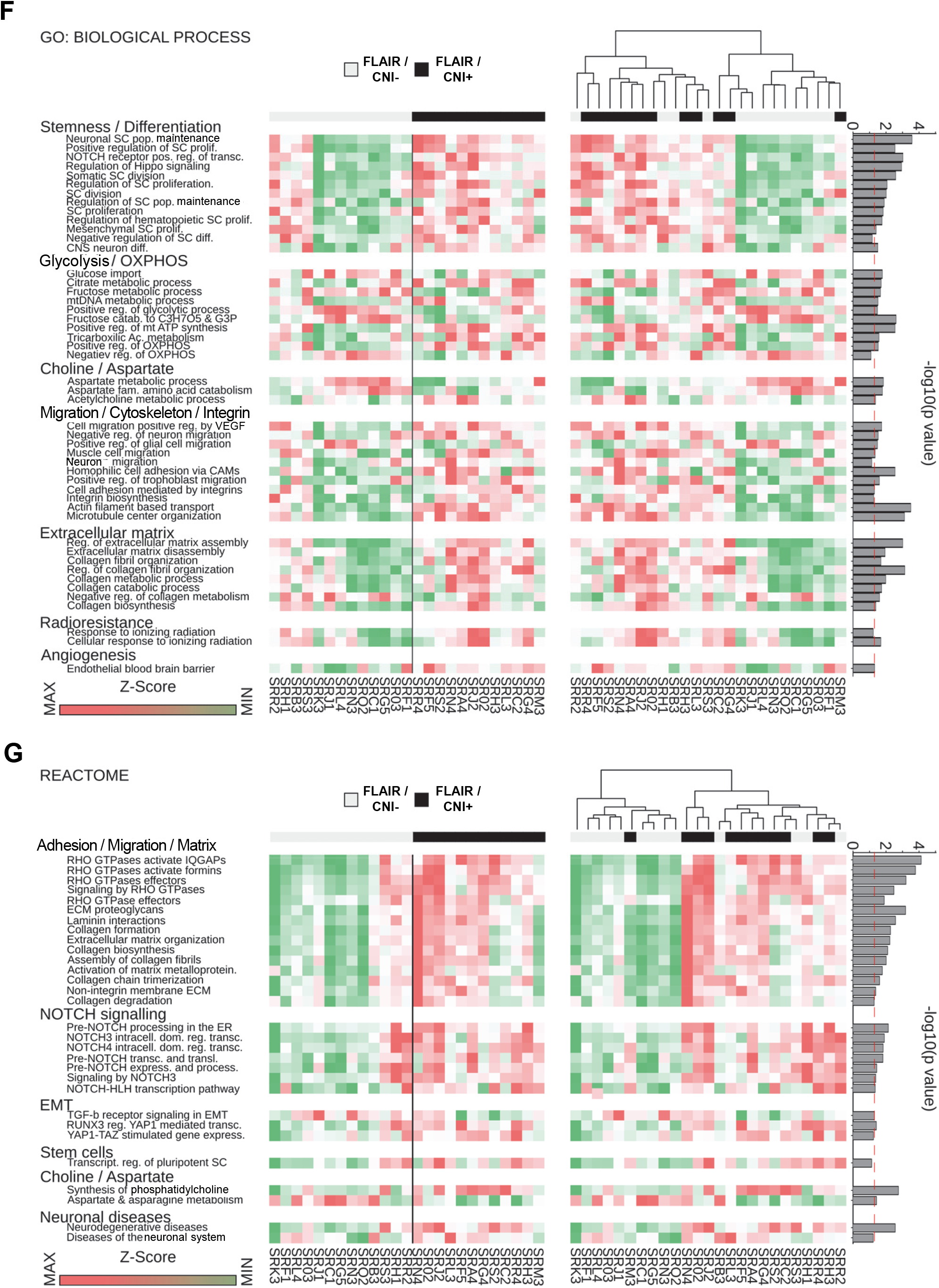
RNAseq analyses revealed that FLAIR/CNI+ biopsies were enriched in gene signatures related to stemness/EMT, migration/adhesion and cell cycle/DNA repair pathways compared to FLAIR/CNI+ tumor samples. **(F-G)** Gene Ontology (GO) Biological Processes **(F)** and REACTOME **(G)** pathway enrichment analyses were performed on FLAIR/CNI− vs FLAIR/CNI+ biopsies and showed the statistically significant differential expression pattern of specified gene signatures through both supervised (left panel) and unsupervised (right panel) analyses.

Beyond this molecular classification, from principal-component analysis (PCA) of the expression data generated from the 4 tumor groups, we found a clear separation of tumor samples from CE and FLAIR samples along the 1^st^ principal component PC1 (accounting for 20.06 % of the total variance), while CE/CNI− and CE/CNI+ clusters were globally overlapping along PC1 and PC2 (2d principal component, accounting for 10.61 % of the total variance) (**Figure 6B**). Moreover, we noticed that 7/13 FLAIR/CNI− samples segregated from the three other groups along PC1. When only FLAIR/CNI− and FLAIR/CNI+ biopsies were taken into account through PCA, we observed more clearly this separation along PC1, with three different clusters: 7 FLAIR/CNI− biopsies on a side, 3 FLAIR/CNI+ samples on the other side and a common cluster between the two (**Figure 6B**). Then PCA suggests that RNAseq analysis may reveal 2 levels of differential expression: FLAIR vs CE, irrespective of CNI pattern, and FLAIR/CNI− vs FAIR/CNI+. A differential expression analysis was subsequently carried out through DESeq2 and MA plots were generated, based on the thresholds of an adjusted q-value < 0.05 and a log2 fold-change ≥ 2. These MA plots revealed that no differential gene expression could be showed either between CE/CNI− and CE/CNI+ subgroups or between CNI− and CNI+ groups, irrespective of the FLAIR/CE origin. However, 4003 differential expressed genes were statistically highlighted between FLAIR and CE groups and 565 between FLAIR/CNI− and FLAIR/CNI+ subgroups (**Figure 6C**).

In order to further explore the differential pattern in FLAIR area between CNI− and CNI+ biopsies, KEGG and HALLMARK pathway enrichment analyses were also performed, which revealed that differentially expressed genes were mainly involved in DNA repair, cell cycle and DNA/RNA-related processes, adhesion/migration and extracellular matrix (ECM)-related pathways, Angiogenesis and stemness/EMT (Epithelial to Mesenchymal Transition), with a clear enrichment of all these pathways in FLAIR/CNI+ biopsies (**Figure 6D**). When focusing on some specific KEGG pathways - Cell Cycle/P53 signaling pathways and ECM Receptor-interaction/Focal Adhesion (FA) - we could observe a strong overexpression of major regulator genes in FLAIR/CNI+ biopsies. For example, considering Cell cycle/P53, *CDK1/2/4, PLK1, ATR, CHEK1, TP53*, and *WEE1* were shown to be significantly overexpressed in FLAIR/CNI+ samples. All of these genes were demonstrated to have major roles in GB progression and progression (**Figure 6E**). By looking at FA/ECM-R interaction pathways, many genes coding for matrix components (Collagen, Laminin, Fibronectin, Tenascin…), integrins (*ITGA1/A3/A4/A7*) and Rho-GTPases activating pathways (*VAV1/3*) were overexpressed in FLAIR/CNI+ areas compared to FLAIR/CNI− (**Figure 6E**). In order to refine this enrichment analysis and to observe in detail which biological processes were differentially represented between these 2 groups, we also conducted REACTOME and Gene Ontology Biological Processes (GO BP) pathway enrichment analyses (**Figure 6F-G**). Supervised (FLAIR/CNI− vs FLAIR/CNI+) and unsupervised analyses revealed a clear and significant enrichment of stemness/EMT-related pathways (NOTCH and Hippo pathways notably) and adhesion/migration/ECM-interaction (relative among others to integrins, Rho-GTPases, collagen and laminin). Moreover, concerning metabolic pathways, we noticed an enrichment of choline-related pathways and a depletion of Aspartate metabolic pathways in FLAIR/CNI+, in accordance with the MRSI parameters established to specifically collect low and high Choline/NAA Index (CNI) areas within tumor and peritumor area (**Figure 6F-G**). Considering other metabolic pathways, we have also observed an important enrichment of mitochondria-related bioenergetic pathways in FLAIR/CNI+ biopsies associated to a lower representation of glycolytic pathways (**Figure 6F**).

As our biological and preclinical results demonstrated a significant increase of GIC subpopulation and GIC-related properties in FLAIR/CNI+ tumor areas compared to FLAIR/CNI−, which was confirmed at the molecular level through Hallmark (**Figure 6D-F**), GO BP and REACTOME (**Figure 6F-G**) pathway enrichment analyses, we also set-up some specific gene signatures to study the enrichment of each areas for either GIC subpopulation or neural-differentiated lineages (Oligodendrocytes, Neurons and Astrocytes), based on our previous papers on the GIC compartment (24, 28) and the related literature (2) (**Figure 7A and supplemental Table 8**). We observed a significant enrichment of the stem-specific signature in FLAIR/CNI+ biopsies and, on the contrary, an enrichment trend for Oligodendrocytic (*p = 0*.*065*) and Neuronal (*p = 0*.*073*) signatures in FLAIR/CNI− zones, showing a strong increase of the undifferentiated/differentiated balance in FLAIR/CNI+ areas compared to FLAIR/CNI− (**Figure 7A**). Of note, no significant differences could be seen for CE/CNI− vs CE/CNI+ or CNI− vs CNI+ subgroups. However, we found a significant decrease of all three differentiation signatures in FLAIR samples compared to CE biopsies. At the gene level, based on the RNAseq results, we indeed observe an important upregulation of numerous specific GIC-related genes (*SOX2, NES, NOTCH1, NG2, ITGA7, EZH2, BIRC5*…) and a global decrease of oligodendrocytic genes (*CNP, OMG, MAL, MBP, OLIG1*…) in FLAIR/CNI+ samples (**Figure 7B and supplemental Figure 5A**). Several astrocytic (*S100B, NDRG2*…) and neuronal (NEFM, SNCA) genes were also decreased in FLAIR/CNI+ biopsies but no clear trend could be highlighted (**supplemental Figure 5B-C**). Of note, when we consider GIC, Oligodendrocytic and Astrocytic markers, it appears that the global expression levels in FLAIR/CNI+ biopsies are quite close to those found in CE/CNI− or CE/CNI+ samples, with no significant differences between the latter two groups (**Figure 7B and supplemental Figure 5**).

**Figure 7.**
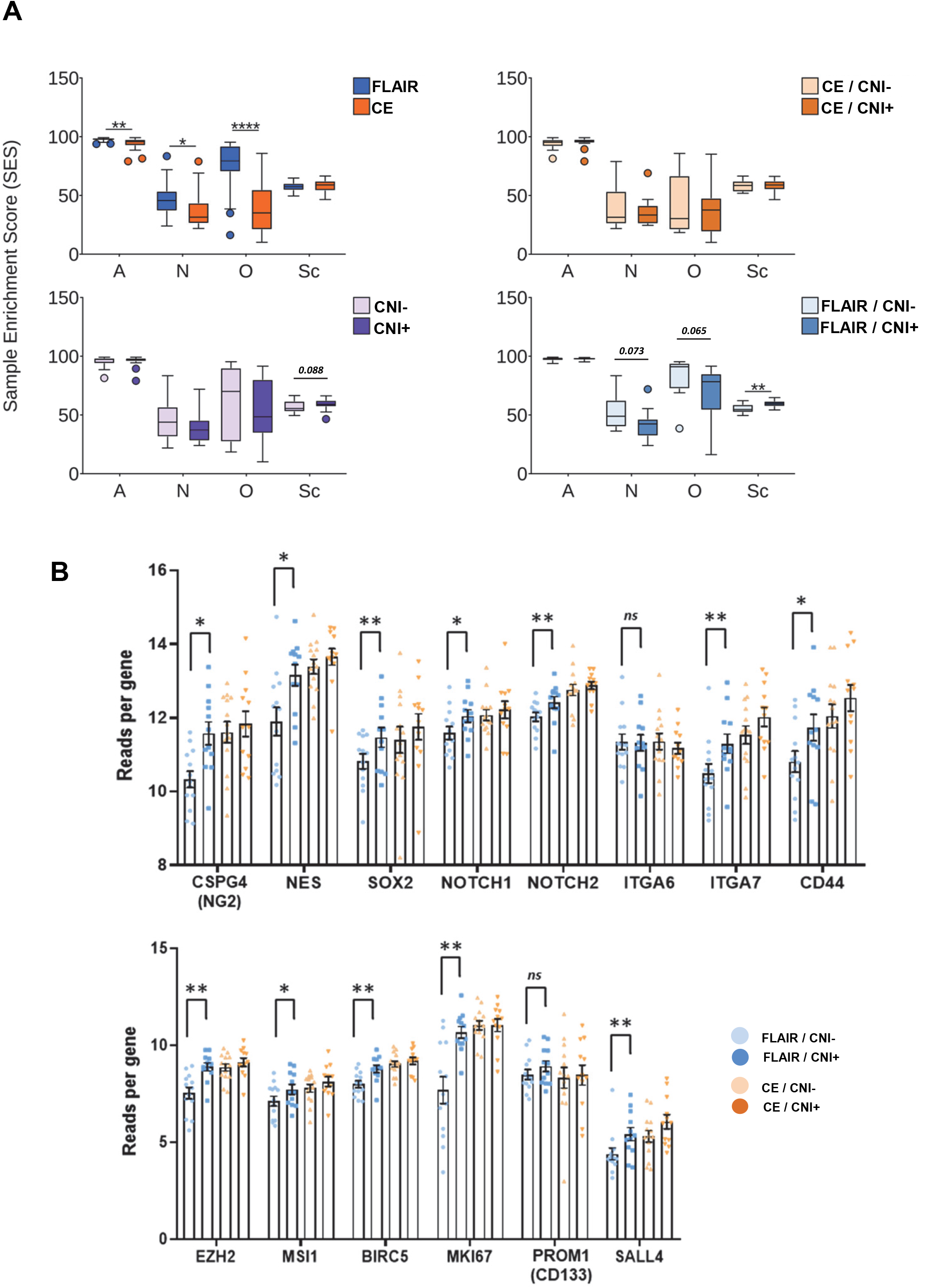
RNAseq-based analyses revealed a differential balance between stemness and differentiation-associated gene signatures between FLAIR vs CE samples and between FLAIR/CNI+ vs FLAIR/CNI− biopsies. RNAseq analyses were conducted on the 51 available tumor biopsies obtained from 16 GB patients and segregated into 4 different groups (FLAIR/CNI−; FLAIR/CNI+; CE/CNI−; CE/CNI+). **(A)** Sample Enrichment Score (SES) were calculated for specifically designed gene signatures: Astrocytes (A), Neurons (N), Oligodendrocytes(O) and Stem Cells (Sc) for the indicated biopsy groups. Boxplot diagrams of the median SES were shown. **(B)** Expression levels for specific GIC-associated genes in the 4 indicated biopsy subgroups were shown as means ± SEM. *\* p ≤ 0*.*05, ** p ≤ 0*.*01, ns*: not significant.

## DISCUSSION

Due to major intratumoral heterogeneity and high cellular plasticity, GB is one of the most difficult solid tumors to manage and treat. As recurrence is almost systematic, determining the tumor prognosis, the response to radio/chemotherapy, the site of recurrence and the degree of tumor invasion are crucial challenges. For more than two decades, MRSI combined to conventional MRI has allowed a better understanding of GB heterogeneity through the determination of some key metabolites at tumor and peritumoral levels (5, 29). Indeed, specific metabolites (such as Chol, NAA, lactate (Lac), lipids, myoinositol (mIns)) or metabolite ratios (such as Cho/NAA, Cho/Cr (creatine) or Lac/NAA) have been shown to improve assessment of GB diagnosis, progression, prognosis and response to treatment, notably to RT. More specifically, increased Cho/NAA index (CNI) and/or Cho/Cr ratio showed great benefit in discriminating glioma grades (30), tumor progression versus RT-induced changes (pseudoprogression or radionecrosis) (5) and in highlighting GB invasion front (17). In addition, CNI was proved to be useful after post-operative RT to assess the prognosis of GB at recurrence, since CNI increase during and/or after RT was associated with early GB progression and bad prognosis (31), notably within the peritumoral/FLAIR edema (12). In line with this, we previously showed that metabolically active CNI>2 regions in both CE and FLAIR areas of pre-RT scans were predictive of the site of relapse after RT (6).

Considering these clinic specificities of CNI+ areas, we designed the STEMRI trial to understand what kind of cellular heterogeneity could sustain such metabolic particularities within the tumor and lead to these localized recurrence zones. We notably focused on the role of GIC since this stem-like cell population was demonstrated to largely mediate recurrence after radiochemotherapy. Here, we demonstrate that FLAIR/CNI+ biopsies contained twice larger amount of GIC subpopulation compared to FLAIR/CNI− samples (median BGC: 6 *vs* 12%) and give rise, in restrictive stem medium, to specific GIC-enriched NS structures faster than FLAIR/CNI− biopsies (median TTN: 13 *vs* 21 days) (**Figure 3, supplemental Figures 1-2 and Tables 1–2**). These FLAIR/CNI+ GIC analyzed by FACS just after surgery seem to overexpress stem markers only quite marginally compared to their CNI− counterparts. These features are only observed in FLAIR area, since CE biopsies, either CNI− or CNI+, possess not significantly different BGC (16.5% and 22.7%, respectively) and TTN (9.5 and 8 days, respectively). Of note, despite a higher content of GIC and a slight overexpression of several stem markers (such as CD44, Nestin or Sox2) in CE biopsies compared to FLAIR samples (median BGC = 12.1% in FLAIR/CNI+), the TTN in both CE/CNI− and CE/CNI+ is relatively similar to TTN in FLAIR/CNI+ samples (13 days). It could be hypothesized that when a threshold BGC value is raised in the biopsy, the delay to generate NS in vitro may be approximately the same.

At the molecular level, RNAseq analysis confirmed these biological observations, with a differential expression analysis revealing major differences in gene expression only between FLAIR/CNI− and FLAIR/CNI+ samples and between CE and FLAIR biopsies. Focusing on FLAIR samples, we demonstrated that CNI+ samples present an enrichment in stemness/EMT-associated pathways, either through the use of GO BP/REACTOME/HALLMARK related signatures (**Figure 6F**) or through the determination of SES (Sample Enrichment Score) for a self-designed dedicated GIC signature (**Figure 7A**). Among these signatures, pathways related to NOTCH, MYC, HEDGEHOG and Hippo/YAP/TAZ appeared to be enriched in FLAIR/CNI+ samples. All these pathways were shown to be major actors to sustain stemness in GIC (32–36). In addition, in FLAIR/CNI+ biopsies, many stem-related genes were significantly overexpressed (**Figure 7B**) and among them SOX2, which was previously showed to be increased at the protein level in high CNI tumor areas in GB (20). On the opposite, we showed that oligodendrocytic and neuronal differentiation signatures and related genes were enriched in FLAIR/CNI− biopsies.

Besides this stem/differentiation balance, our GO BP analyzes also reveal a significant enrichment in FLAIR/CNI+ zones for oxidative mitochondrial metabolic pathways in detriment of glycolytic processes. In this respect, the energetic metabolism in GIC appears to be very plastic and to rely partly on oxidative phosphorylation, and not predominantly on glycolysis like differentiated GB cells (37–39). Moreover, we showed that FLAIR/CNI+ bulk samples overexpressed numerous gene signatures (KEGG, GO BP and Reactome) related to cell adhesion, migration and interaction with extracellular matrix. Interestingly, it has been described that GSC have an increased ability to adhere, migrate and invade compared to their differentiated counterparts (40, 41), but also that those capacities may vary according to the anatomical location of the biopsy, since GIC generated from the GB peritumoral area displayed a higher migratory and invasive phenotype compared to GIC isolated from the tumor mass (42). In a recent study associating MRSI and RNAseq of tumor bulks both in GB patients and in paired patient-derived orthotopic xenografts (PDOX) (43), it was pointed out (i) that there was a good concordance between metabolite profiles recorded by MRSI in patients and the metabolite profiles observed during PDOX growth and (ii) that this tumor growth and related metabolic profiles (notably high choline-containing compounds and low NAA), both in human and PDOX, were associated at the molecular level with an enrichment in signatures dedicated to adhesion, invasion and extracellular matrix, consistent with our results. All these observations lead us to hypothesize that GIC-enriched NS isolated from FLAIR/CNI+ areas might exhibit higher migratory/invasive and metabolic heterogeneity that could be monitored by at least RNAseq and MRSI on cells (44) and in xenografted models (21, 43). These results could then be translated and validated at the clinical level through their specific therapeutic targeting to *in fine* improve radiochemotherapeutic response.

In complement to the overexpression of these different pathways in FLAIR/CNI+ bulk samples, we have shown that the NS coming from FLAIR/CNI+ biopsies were generated *in vitro* faster than their CNI− counterparts (shorter TTN) but also that these NS were more tumorigenic in orthotopically-xenografted mice (**Figures 4A**). Although no significant difference can be highlighted between stem markers at day 0 in FLAIR/CNI− vs FLAIR/CNI+ GIC, this could suggest, since tumorigenicity is a main characteristic of cancer stem-like cells (3), that primary NS cell lines generated from FLAIR/CNI+ areas may possess more robust stem cell-like traits than FLAIR/CNI− NS. Of note, one of the characteristics of GIC compared to more differentiated GB cells is their strong radioresistance. However, we failed to observe a higher MID in FLAIR/CNI+ NS, despite an enrichment in GO BP signatures associated with response to RT in FLAIR/CNI+ bulk biopsies (**Figure 6F**). One of the main challenges for future studies will be to explore more specifically stem-related properties of both FLAIR/CNI− and FLAIR/CNI+ GIC, particularly through RNAseq analyses and clonogenic limiting dilution assays, and to decipher whether or not CNI+ NS could better resist to chemo or radiotherapy.

All these biological arguments in favor of an increased stem population in FLAIR/CNI+ GB areas, combined with the fact that these tumor zones are associated in clinic with early progression, bad prognosis and recurrence site prediction, strongly suggest that it might be a correlation between our two biomarkers (BGC and TTN) and the clinical outcome of GB patients. We effectively observed that BGC% at day 0 in FLAIR/CNI+ biopsy is inversely correlated with OS and PFS. In the same way, the TTN-derived parameter measuring the ability of FLAIR/CNI+ biopsy to generate primary NS at d14 (PN14) is also correlated with OS and PFS. We can therefore conclude that in FLAIR/CNI+ tumor areas, the determination of these two GIC-related markers can predict patient outcome. This may have a major importance in clinic for GB treatment to delay recurrence after surgery and radiochemotherapy. Indeed, the gold standard surgery for GB is presently the ‘maximal safe resection’ based on the maximal removal of CE tumor zones, without FLAIR-positive peritumoral area resection. It was previously shown that larger extent of CE gross total resection (notably in the range of 80 to 100%) is associated with improved patient outcome (45). But recently supramaximal resection of both CE and FLAIR tumor zones was also experimented in large cohorts of GB patient. These trials demonstrated that minimal residual extent of non-CE tumor after surgery or higher removal extent of FLAIR-positive edematous infiltrative tumor area could translate into patient outcome benefits (45–47). However, the most important limitation of this new surgical procedure is the possible manifestation of new postoperative neurological deficits linked to the supramaximal resection of FLAIR tumor zones, which could significantly hamper the survival benefit seen in these patients (48). In the light of these new clinical data and our present results, we can hypothesize that restricting the resection of the edematous and infiltrated non-CE zone to the FLAIR areas associated with GIC enrichment (i.e. here the CNI+ zones determined by MRSI) could certainly reduce the risk of postoperative neurological deficits and thus significantly increase patient outcome. Moreover, beyond MRSI, we have recently demonstrated through preoperative perfusion and diffusion MRI that increased Cerebral Blood Volume (rCBV) and low Apparent Diffusion Coefficient (ADC) could correlate respectively with high BGC% and shorter TTN in FLAIR biopsies (49). Then, combining MRSI/MRI data with multimodal MRI (notably perfusion and diffusion MRI) could also help to target more precisely those particular FLAIR areas associated with higher recurrence and tumorigenesis and reduce neurological side-effects.

Finally, by highlighting in CNI+ areas several major pathways that we have previously shown to be involved in resistance to RT, as EMT-related pathways, DNA repair-linked pathways or pathways associated to adhesion/migration/ECM-interaction (33, 50), these results also suggest that, beside using MRSI for guided surgery and radiotherapy target determination, association of inhibitors of these particular pathways in association with radio-chemotherapy could also increase the efficacy of these treatments in the management of patients with GB.

## METHODS

### Patients

#### Study approval

This trial was reviewed and approved by the institutional review board, the French ethics committee on 30/05/2012 (registration number 2012-A00585-38) and the French Drug Administration (ANSM) on 15/06/2012 (registration number B120639-40). All subjects signed a written informed consent form (**Figure 1**).

#### Study design

Between May 2013 and June 2017, 21 patients with newly diagnosed GB have been enrolled in a prospective biomedical study of interventional type. This pilot study (ClinicalTrials.gov NCT01872221), combining a metabolic imaging approach (Proton Magnetic Resonance Spectroscopy or ^1^H-MRSI) and a biological one, have been performed in GB patients to determine whether a particular MRSI marker of aggressiveness (CNI>2, hereafter referred to as CNI+) can be associated with specific biological patterns as regards to GIC content.

In the first part of the study, patients with radiological criteria of GB amenable to surgical resection have been selected by a preoperative committee and then included **(Figure 1)**. Pre-operative multimodal MRI/MRSI scans have been done (CHU Purpan, Toulouse, France) and all data acquired have been integrated in the image-guided surgical device (i.e. neuronavigation system) to be used intraoperatively (cf. Patient’s imaging paragraph for details). During tumor resection (CHU Purpan), tissue samples have been individualized, based on their multimodal imaging characteristics and processed for biological analysis (as described below). Of note, post-operative MRI were realized in the 72h hours following surgery and also before radiotherapy for treatment planning (at day 30 post-surgery). In the second part, patients were treated after surgery by the standard radio-chemotherapy Stupp (51) protocol (Institut Claudius Regaud, Toulouse, France) and then followed starting one month after the end of the chemo-radiotherapy phase, by clinical evaluation and multimodal MRI including MRSI every 2 months during the first year (CHU Purpan) and then every 3 months until progression **(Figure 1)**.

#### Study objectives

The study primary objective outcome was the capacity of GB resected samples to form GIC-enriched neurospheres (NS) *in vitro* as well as to form invasive brain tumor after orthotopic xenograft of the derived GIC population in nude mice. The *in vitro* capacity was assessed at the CRCT center (Toulouse, France) through several parameters: (i) the time necessary to generate NS after biopsy processing and culturing and (ii) the GIC-enrichment of the resected biopsies just after surgery, determined by GIC-subpopulation quantification through FACS analysis and by the determination of particular molecular stem signatures in resected samples through RNAseq analysis. The *in vivo* capacity was both assessed by the percentage of GIC-implanted mice able to develop a brain tumor and the time to onset of tumor-associated neurological signs.

The secondary objective outcome was the time to progression (surgery to progression) and the overall survival (time from surgery until death or last of follow-up news)

#### Patient’s selection

For the first part of the study, inclusion criteria concerned patients ≥ 18 years old, patients with performance scale ECOG 0-2, patients with life expectancy ≥ 3 months, patients who presented a radiological criteria of GB and who had a surgical indication. Patients should also be affiliated to social security regimen and have voluntarily agreed to participate by giving written informed consent for this first part. For the second part of the study, the additional inclusion criterion was the presence of a histologically confirmed GB. The exclusion criteria for the first part of the study concerned patients who were not allowed to perform an MRI, patients with non-contributive results after spectroscopic exam, pregnant or nursing patients, patients under law protection, patients who presented conditions that would interfere with cooperation with the trial requirements and patients who presented medical, severe or chronic biological or psychiatric conditions not allowing an enrolment in the study, according to the investigator’s opinion. For the second part of the study, the exclusion criterion was receipt of the biological material in the laboratory more than 48 hours after surgery. As a consequence, 16 patients were finally included in the second part of the trial and evaluable for the entire study (**Table 1**).

### Patients’ imaging

#### MRI/MRSI acquisition

As described (49), data were acquired under the neurosurgeon supervision on a 3T MRI system preceding surgery (ACHIEVA dStream, Philips Healthcare). A 32-channel phased-array receive coil was used. Two MRI sequences were then acquired (CHU Purpan). First, the anatomical MRI acquisition protocol included a 3D T1-weighted after 15 ml injection of Gadolinium contrast (TR/TE=8/4ms, FA=8°, matrix=165×241, 240 slices, 1×1×1mm^3^ resolution), a FLAIR (TI/TR/TE=2400/8000/335ms, FA=90°, matrix=200×256, 256 slices, resolution=1×1×1mm^3^) and a turbo-spin echo T2w (TR/TE=4130/80ms, FA=90°, matrix=512×512, 43 slices, resolution=0.5×0.5×3mm^3^). Then, the 2D Spectroscopy acquisition sequence (^1^H-MRSI) was performed using a point-resolved spectroscopy acquisition (PRESS) over 4 slices covering the lesions. Matrix= 8×8, voxel size=1×1×1cm^3^, TR/TE=1000/144ms, two averages. Total acquisition time was around 45-50 min. Field of view was shifted away from brain-air interfaces when necessary.

#### MRI/MRSI data processing

Spectral processing (water subtraction, low-pass filtering, frequency-shift correction, baseline and phase correction, and curve fitting in the frequency domain) and the computation of choline (Cho)-to-N-acetyl-aspartate (NAA) ratio index (CNI) map were performed with the Syngo MR B17 spectroscopy application (Siemens). Data were then processed according to our previous published studies (6, 7, 49).

Biopsy volume is defined by the needle size of 1.8mm. The precision of the location is defined by (i) the spatial registration of the neuronavigation system (4mm), (ii) the stability of the needle during extraction (approximately 2mm), and (iii) the resolution of the anatomical image used for guidance (1mm, leading to a precision of 1/√12mm at 1σ). Finally, the biopsy lies in a volume of 680mm3 at 1σ, or 2040mm^3^ at 3σ, around the estimated location. Starting from the voxel corresponding to the estimated biopsy location, a morphological dilation was applied until the region of interest (ROI) reached 2040 mm3 to take into account these uncertainties. This dilatation was constrained in the manually drawn hyper-FLAIR region.

### Tissue specimen surgical collection

For each patient, pre-operative multimodal MRI was performed and tumor localization was determined. During surgery, within the standard protocol set up in our hospital, a biopsy of the tumor was taken from the contrast enhancement (CE) area, independently of the MRSI parameters, in order to be analyzed by the anatomopathology department for MGMT and IDH status determination.

As part of this clinical trial, as described in our recent study (49), the tumors were manually segmented according two preoperative MRI-visualized components: tumor bulk (CE and necrosis) and tumor edema (FLAIR) (**Figure 2B-C**). Hence tumors were categorized into tumor functional zones and segmentation in each zone (CE or FLAIR) was performed according to the CNI >2 or <2 regions (CNI+ or CNI−, respectively). The segmented tumor volumes, tractography, and computed maps were transferred to the neuronavigation planning system BrainLab iPlan, and the operating room BrainLab VectorVision Neuronavigation System (Nnav). Relative to surgery and biopsy extraction, in order to minimize brain shift, two biopsies were performed prior to resection according to the CNI >2 and <2 in the peritumoral edema regions using intraoperative MRI guidance. Resection was then performed according to the surgical standards of maximum safe resection of CE and more if possible, using neuronavigation and refined to take into account electro-stimulation. During this CE resection, biopsies arising from CNI>2 and <2 areas were performed.

To sum up, for each patient, when possible according to surgical constraints and mapping results, it was planned to obtain a maximum of four different types of biopsies: FLAIR/CNI− (group 1); FLAIR/CNI+ (group 2); CE/CNI− (group 3); CE/CNI+ (group 4). The correspondences between patients and collected samples are detailed in **Table 2**.

### Tissue specimen processing and culture

After extraction, biopsies were processed using our established protocol (28) and a small fraction of the sample was directly analyzed by flow cytometry to study the percentage of the GIC subpopulation at day 0 (Biopsy GIC Content or BGC%) over the total number of cells, as previously described (28). Next, another small fraction of each biopsy was set aside for RNAseq analyses. Finally, remaining cells were cultured as GIC-enriched neurospheres (NS) in stem cell medium, as described in (28). In these restrictive conditions, only GIC can survive and form NS. All cultivated samples were observed daily under a microscope to check for the appearance of NS and thus report the Time to NS (TTN) formation in days. Of note, the BGC% can be considered as a marker of malign stem-like cells’ infiltration in the tumor biopsy and the TTN as a surrogate marker of tumor cell aggressiveness as shown in patients (52) and in xenografted mice (53).

### MGMT and IDH status

IDH1/2 mutation and MGMT promoter methylation status were determined in all patients. To perform DNA extraction from formalin-fixed paraffin-embedded (FFPE) tissues, Maxwell® RSC DNA FFPE Kit (Promega) was used. DNA was quantified by fluorimetric assay with DNA Qubit, Broad Range (BR) Kit (Thermo Fisher Scientific) and purity was checked by NanoDrop ND-100 (Thermo Fisher Scientific).

We used pyrosequencing technology (PyroMark Q24, Qiagen) for real-time, sequence-based detection and quantification of sequence variants IDH1/2. Prior to Pyrosequencing reaction, we made biotinylated-PCR reaction using Pyromark PCR kit (Qiagen). Concerning biotinylated-PCR primers, for IDH1 (NM_005896), we targeted the p.R132 codon and used the following pair of primers to generate a 74pb amplicon size (IDH1-F-Biot: 5’-Biot-GGCTTGTGAGTGGATGGGTA-3’, IDH1-R: 5’-GCCAACATGACTTACTTGATCC-3’). For IDH2 (NM_002168), we targeted the p.R172 codon and used the following pair of primers to generate a 82pb amplicon size (IDH2-F1: 5’-ATCCCACGCCTAGTCCCT-3’; IDH2-R1-Biot: 5’-Biot-CTCCACCCTGGCCTACCT-3’).

Concerning the sequencing primers, since a single-stranded DNA template is required for pyrosequencing, the second strand must be removed. The pyrosequencing reaction have been performed on the purified biotinylated DNA template strand using the following sequencing primers: IDH1-S(R): 5’-GACTTACTTGATCCCCATA-3, IDH2-S1(F): 5’-AAGCCCATCACCATT-3’. As pyrosequencing uses sequential nucleotide injection, the nucleotides dispensation order allows quantification at all the chosen variant positions (IDH1: GAGCATCGATCTGACTA; IDH2: CGTCATGCAC). Finally, the PyroMark Q24 Software 2.0 was used in AQ mode for pyrogram analyse of listed IDH1 variants: p.R132H; p.R132C; p.R132L; p.R132S and p.R132G. For this we used 2 different analysis sequences (AGCATGAHGACCTAT and AGCATGACNACCTAT) in order to call and detect the targeted mutation. To detect the listed IDH2 variants: p.R172M; p.R172K; p.R172W and p.R172G, we used 2 different sequences (GGCADGCACGCCCAT and GGCDGGCACGCCCAT).

MGMT promoter methylation was assessed by pyrosequencing. Prior to pyrosequencing, Bisulfite conversion was performed with EZ DNA Methylation Kit (ZYMO RESEARCH) according to the supplier’s recommendations as well as all the steps leading to MGMT pyrosequencing with the MGMT Pyro kit (Qiagen). MGMT Pyro Kit is used for quantitative measurement of methylation of four CpG sites in exon 1 of the human MGMT gene.

### RNAseq analysis of tumor bulk biopsies

RNA were isolated from frozen tissue material. RNeasy lipid Tissue Mini Kit (Qiagen) was used, combined with Qiazol Reagent (Qiagen) to perform RNA extraction. RNA have been quantified by fluorimetric assay with RNA Qubit Broad Range Kit (Thermo Fisher Scientific) and purity was checked by NanoDrop ND-100 (Thermo Fisher Scientific). The RNA Quality have been checked with Bioanalyser 2100 (Agilent Technology) to get RIN (RNA integrity number) and to check for the absence of genomic DNA. 100ng of total RNA have been be used for library preparation. NGS-based RNA sequencing have been performed by Helixio (Clermont-Ferrand, France) using the Illumina TruSeq Stranded Total RNA sample preparation with Ribo-Zero Gold depletion (Illumina) on a Nextseq 550 instrument according to the manufacturer’s instructions (Illumina).

FastQ files were generated via llumina bcl2fastq2 (version 2.20.0.422), them verified for quality (FastQC/MultiQC), aligned and quantified using RSEM (RSEM 1.3.0, bowtie2-2.3.4) and the *Homo sapiens* transcriptome reference GRCh38 (v97) (54). Differential analysis of RNAseq data (counts) at the gene level was performed using the DESeq2 (R package) with the recommended workflow (55). Adjusted *P* values (q values) were obtained as default by DESeq2 to control the global FDR across all comparisons. Genes were considered differentially expressed if they had an adjusted q value < 0.05 and a log2 fold-change ≥ 2. Pathway enrichment analyses were performed through successive analysis of the KEGG (C2_CP_KEGG), HALLMARK (H), REACTOME (C2_CP_REACTOME) and Gene Ontology - Biological Process (C5_GO_BP) databases with the AutoCompare Sample Enrichment Score (SES) tool. Finally, the obtained scores by samples for each group (FLAIR/CNI−; FLAIR/CNI+; CE/CNI−; CE/CNI+; CNI−; CNI+; CE; FLAIR) were compared using a T-test. Significant signatures were those with a *P* value < 0.05. The different databases and tools are available from Fred’s Softwares at https://sites.google.com/site/fredsoftwares/products (56).

Molecular subtype of each biopsy was determined according to either Verhaak (25) or Wang (26) classification. For Verhaak classification, Simple GB Subclassifier online tool (www.semel.ucla.edu/coppola-lab/simple-glioblastoma-subclassifier) was used to classify samples into 4 molecular subgroups (Ne: Neural; Pn: proneural; Mes: Mesenchymal; Cl: Classical). While for Wang classification, the Gliovis data portal and the SubtypeME tool (http://gliovis.bioinfo.cnio.es) (27) were used to classify biopsies into 3 tumor-specific subtypes: Pn, Mes and CL.

### Flow cytometry

Direct immunofluorescence assay was performed by FACS as described (24, 28). The antibodies used were A2B5-APC and O4-PE (Miltenyi Biotech), CD133/2-PE, CD15-PerCP, NOTCH1-PerCP, NANOG-PE, Nestin-fluorescein, ITGβ8-APC and Sox2-APC (R&D Systems); GFAP-PE (Millipore,); TUJ1-AlexaFluor 488, KI67-PECy7 and CD44-PECy7 (BD Biosciences) and ITGα6-PE (Thermo Fisher Scientific). For marker expression results, specific fluorescence index (SFI) and gating strategy were previously described (28).

### Clonogenic irradiation assays

NS from different patients and localizations were seeded in 96-well plates (100 cells/wells, 16 wells per condition). After 24 h, cells were treated or not with different doses of gamma rays (2 to 10 Gy). Then, 8–15 days post-IR, the number of NS/well with more than 50 cells was measured. The best fit survival curve was generated according to the linear quadratic model and the mean inactivation dose (MID) calculated, as already detailed (57).

### Orthotopic xenografts

Nude mice were used in accordance to a protocol (APAFlS#7660-2016110818123504 v2) reviewed and approved by the Institutional Animal Care and Use Committee of Région Midi-Pyrénées (France). Orthotopic human GB xenografts were established in 4-6 weeks-old female nude mice (Janvier Labs) as previously described (28). Briefly, mice received a stereotaxically guided injection of 2.5×10^5^ cells resuspended in 5µl of DMEM-F12. Survival curves were established and mice were sacrificed at the appearance of neurological signs. Excised brains were collected for subsequent immunohistochemistry analysis.

### Immunohistochemistry (IHC)

IHC was performed, as previously described (24), on paraffin-embedded sections (5 µm) of excised brains of xenografted mice. The primary antibody used was mouse anti-Nestin (Millipore MAB5326)..

### Statistical analysis

As regard to the exploratory nature of this pilot study, no formal sample size calculation was performed and it was planned to include sixteen eligible patients. Data were described using frequencies and percentages for qualitative variables and using median and range or mean and SEM (standard error of the mean) for quantitative variables. Comparisons between pairs of biopsies CNI+ versus CNI− were performed using the Wilcoxon signed rank test for paired data for quantitative variables and the McNemar test for qualitative variables. Overall survival (OS) and Progression-free survival (PFS) were estimated using the Kaplan-Meier method. Univariable analyses were performed using Log-rank test for qualitative variables and Cox proportional hazard model for quantitative variables. All statistical tests were two-sided and a p-value <0.05 was considered as statistically significant. Statistical analysis was performed using Stata software version 16.

## Data Availability

All data produced in the present work are contained in the manuscript, with the exception of the raw RNAseq data, which will be publicly available upon acceptance for publication.

## AUTHOR CONTRIBUTIONS

ECJM, VL and CT conceptualized the study. ECJM, CT, CD and A Lemarié designed the experiments. ECJM coordinated the trial. ECJM and VL recruited patients. ECJM, MM and VL proceesed the clinical data. TF and A Lusque analyzed the data and supervised the statistical and analytical methods used. CD and A. Lemarié processed and analyzed tumor tissues samples. CD, A. Lemarié, PD and FA established and cultured the primary cell lines. AS and YN set up and performed the pathological analyses on tumor samples. JPC and MP analyzed the RNAseq data. A. Lemarié and ECJM wrote the manuscript with comments from all authors.

## ACKNOWLEDGEMENTS

We want to acknowledge the Institut Claudius Regaud and GRICR, RITC foundation and the HTE program from Plan Cancer 2016 (MoGlimaging). We would like to thank Emmanuelle Uro-Coste, Monique Courtade-Saïdi and Solène Evrard at the CHU Toulouse-IUCT-oncopole Anatomopathology service, as well as Aurianne Hagimont for technical advises. We would also like to thank the CRCT Technology Cluster, the CREFRE animal facility in Toulouse, INSERM and the Association pour la Recherche sur les Tumeurs Cérébrales (ARTC).

