## Supplemental Figures and Tables for "The STEMRI trial: magnetic resonance spectroscopy imaging can define tumor areas enriched in glioblastoma-initiating cells"

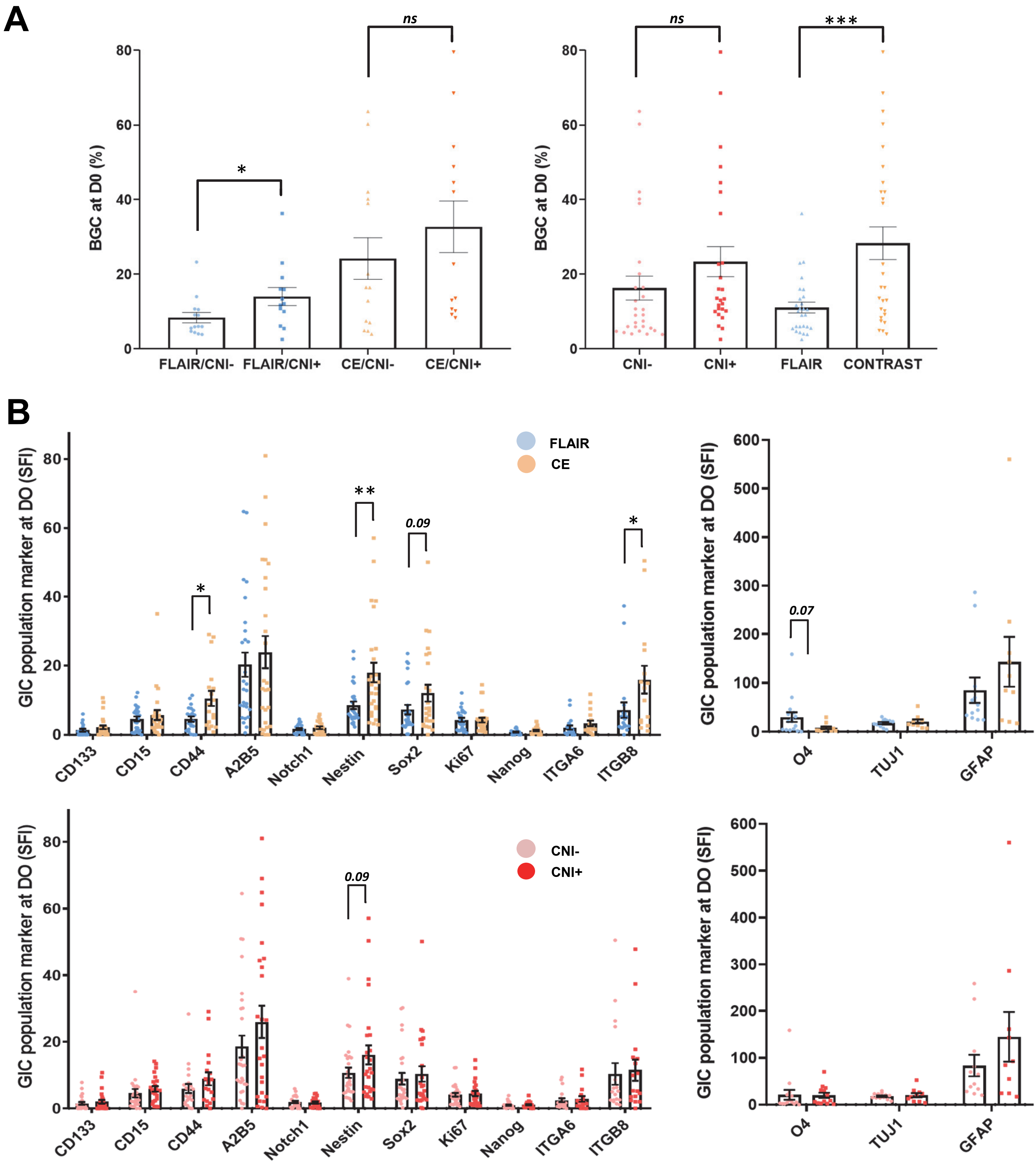


**Supplemental Figure 1. Analysis of Biopsy GIC content and stem/differentiation markers' expression in the different biopsy subgroups at day 0. (A)** Comparative analysis across all biopsy samples of the Biopsy GIC Content at day 0 (D0) after surgery (BGC%), as determined by FACS, either in FLAIR/CNI-, FLAIR/CNI+, CE/CNI- and CE/CNI+ samples (left panel) or in CNI- vs CNI+ or FLAIR vs CE samples (right panel). **(B)** Immunofluorescence analysis performed by FACS for the indicated stem (left panel) and differentiation (right panel) markers in the GIC subpopulation analyzed at D0 either in FLAIR vs CE samples (upper panels) or in CNI- vs CNI+ samples (lower panels). The SFI (Specific Fluorescence Index) was used to evaluate the marker expression level. **(A-B)** Results were expressed as means ± SEM of all analyzed samples. * *$p \leq 0.05$*, ** *$p \leq 0.01$*, *** *$p \leq 0.001$*.

A

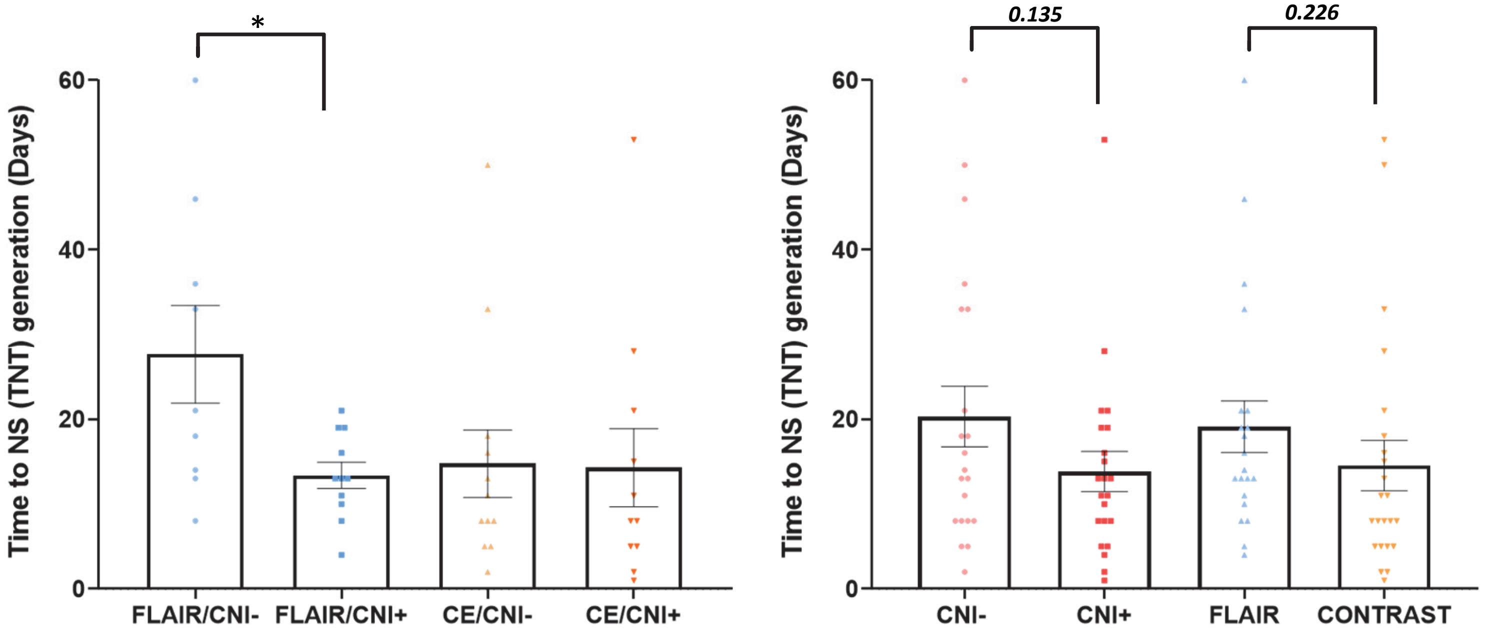


**Supplemental Figure 2. Analysis of both ability and delay to generate neurosphere (NS) from biopsies across the different subgroups.** Comparative analysis across all biopsy samples of the Time to NS (TTN) generation either in FLAIR/CNI-, FLAIR/CNI+, CE/CNI- and CE/CNI+ samples (left panel) or in CNI- vs CNI+ or FLAIR vs CE samples (right panel). Results were expressed as means ± SEM of all analyzed samples. * $p \leq 0.05$.

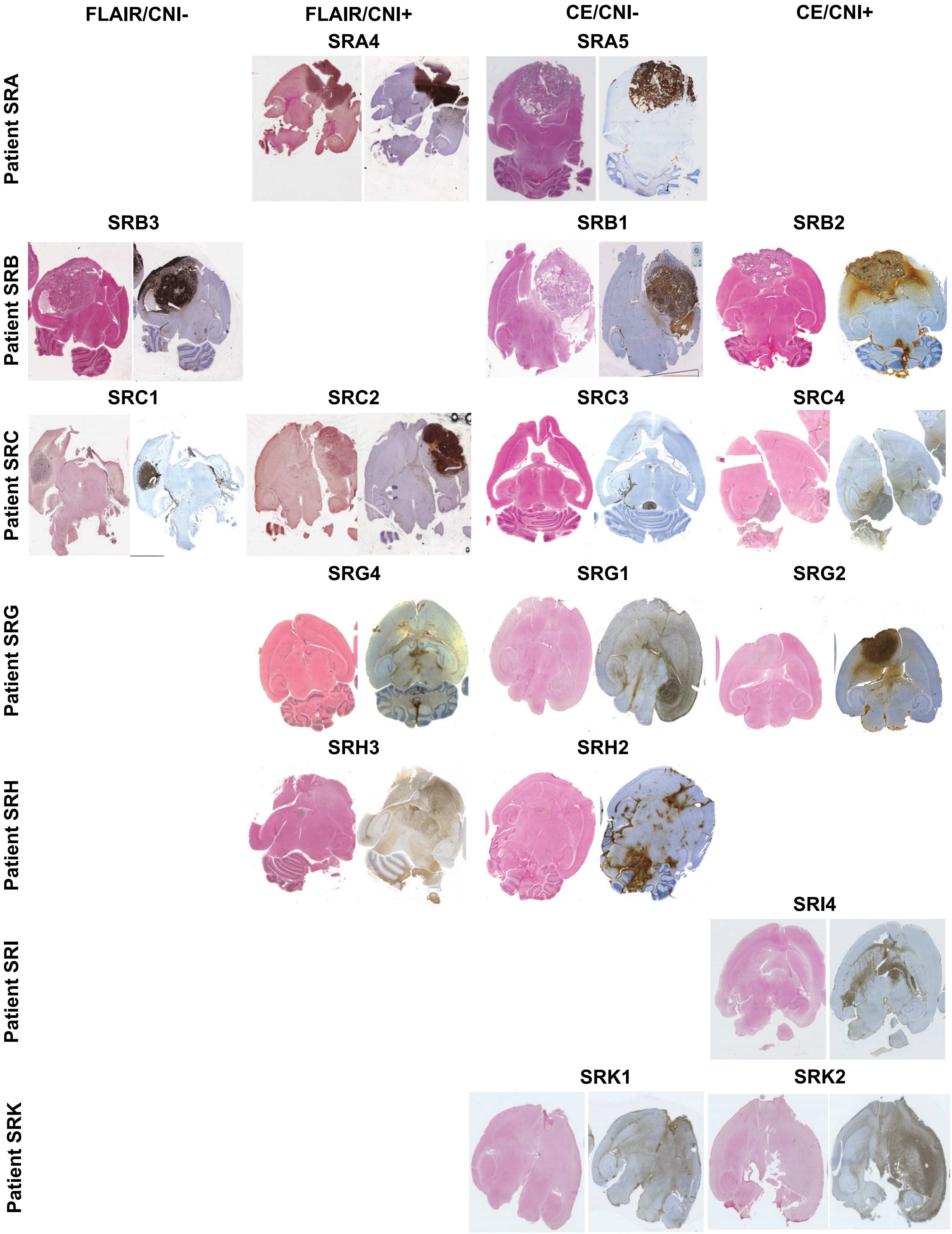

FLAIR/CNI-
FLAIR/CNI+
CE/CNI-
CE/CNI+
Patient SRA
SRA4
SRA5
Patient SRB
SRB3
SRB1
SRB2
Patient SRC
SRC1
SRC2
SRC3
SRC4
Patient SRG
SRG4
SRG1
SRG2
Patient SRH
SRH3
SRH2
Patient SRI
SRI4
Patient SRK
SRK1
SRK2

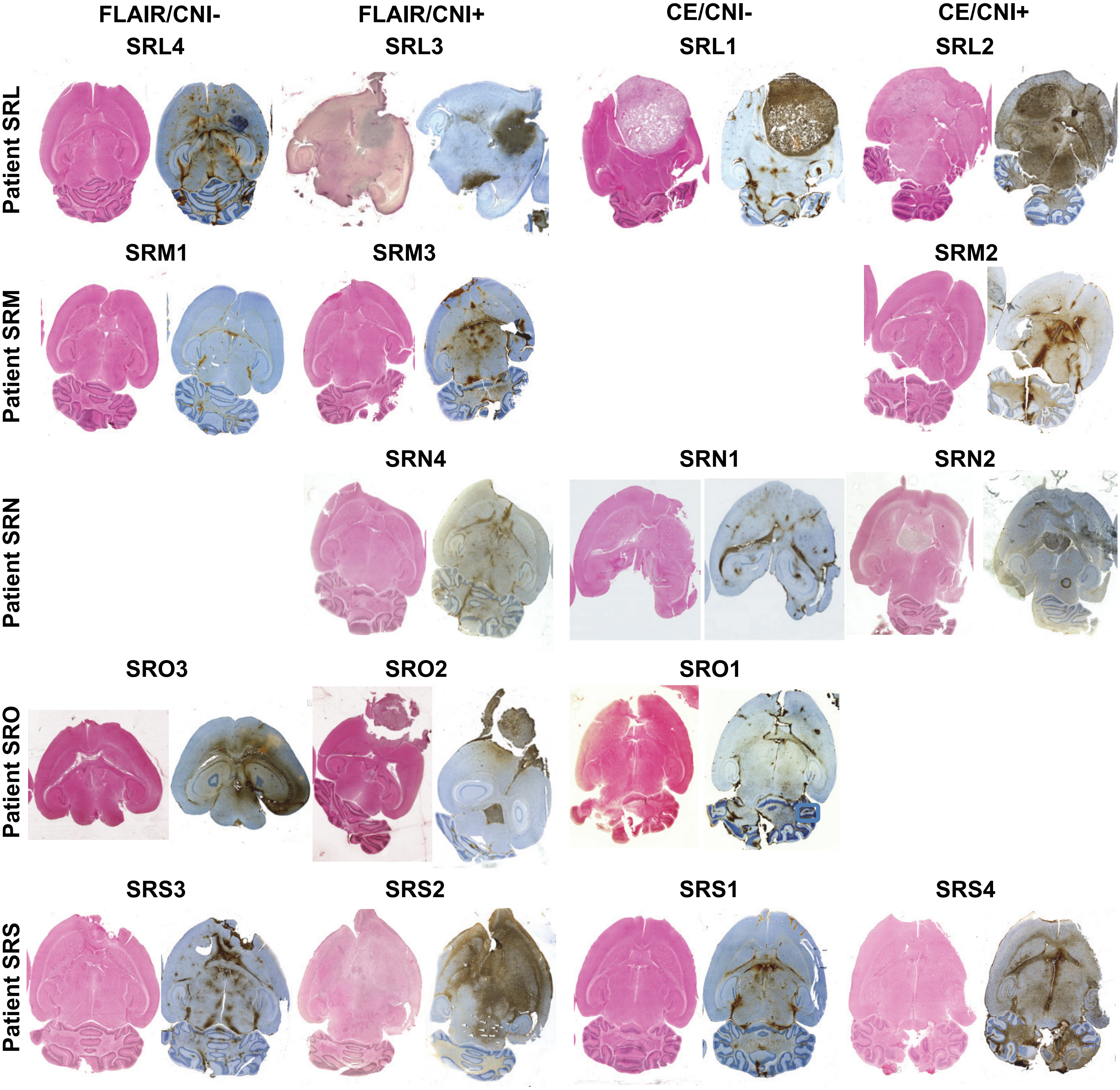


**Supplemental Figure 3. Tumorigenic potential of GIC cell lines isolated from patient biopsies in orthotopically-xenografted nude mice.** For all biopsy subgroups (FLAIR/CNI-, FLAIR/CNI+, CE/CNI- and CE/CNI+), each primary GIC cell line able to generate stable secondary NS (n = 34) was dissociated and then subjected to orthotopic xenografts in nude mice (n = 4-6 mice per NS samples). The tumorigenic potential of these xenografted GIC was assess through hemalun staining (left panels) and Nestin immunostaining (right panels). Shown are some representative scans of Hemalun and Nestin staining of at least 3 implanted mice.

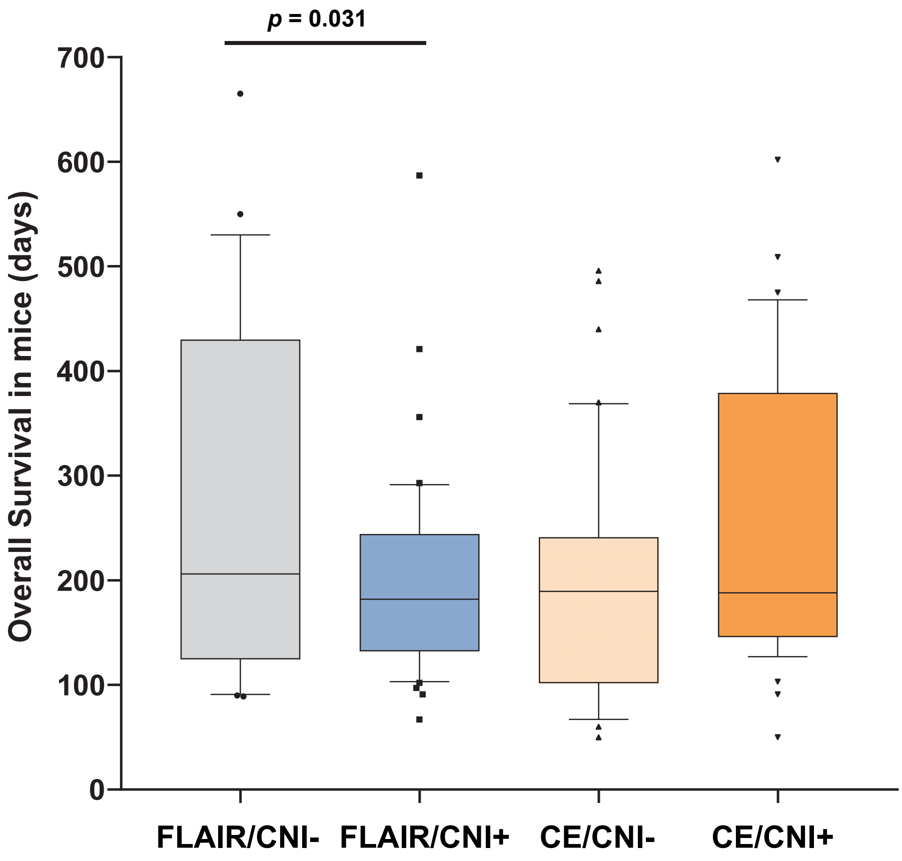


**Supplemental Figure 4. GIC primary cell lines obtained from FLAIR/CNI+ biopsies are associated with increased tumorigenesis in xenografted mice compared to FLAIR/CNI- neurospheres.** For each primary GIC cell lines able to generate stable secondary neurospheres, NS-dissociated cells were subjected to orthotopic xenografts in nude mice (n = 4-6 mice per sample). Overall survival (OS) data were determined for each GIC groups (FLAIR/CNI-; FLAIR/CNI+; CE/CNI-; CE/CNI+) in order to assess their tumorigenic ability. Boxplot diagram of the median OS in mice (in days ± SEM) is depicted.

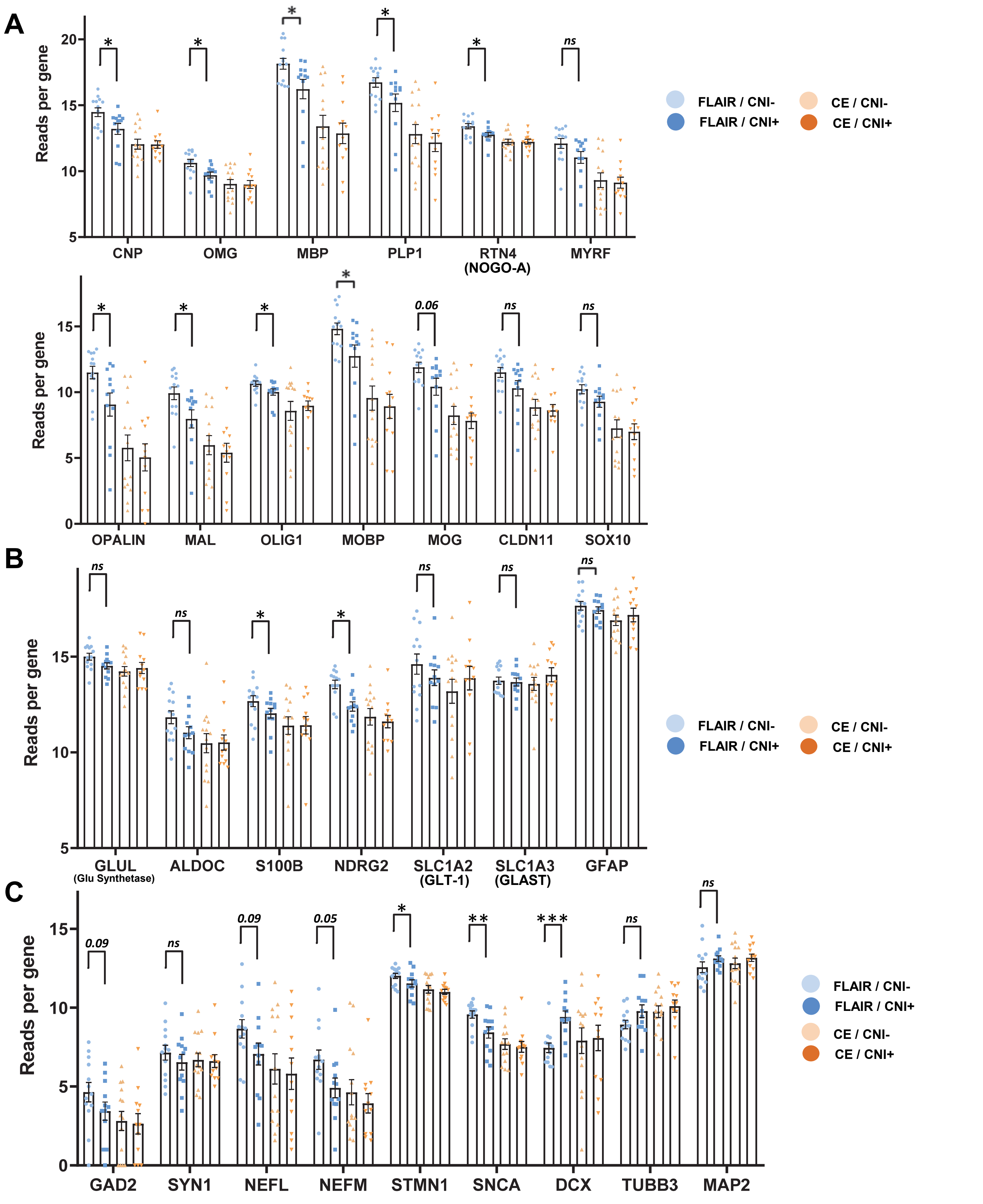


**Supplemental Figure 5. RNAseq-based analysis of the gene expression for several lineage-specific neural differentiation markers across all biopsy subgroups.** RNAseq analyses were conducted on the 51 available tumor biopsies obtained from 16 GB patients and segregated into 4 different groups (FLAIR/CNI-; FLAIR/CNI+; CE/CNI-; CE/CNI+). **(A-C)** Gene expression levels for a panel of markers associated to either oligodendrocytic **(A)**, astrocytic **(B)** or neuronal (**C)** differentiation status in the 4 indicated biopsy subgroups. Results were expressed as means ± SEM of all analyzed samples. * $p \leq 0.05$, ** $p \leq 0.01$, *** $p \leq 0.001$.

| | Biopsy localization | | | |
|---|---|---|---|---|
| | FLAIR/CNI- | FLAIR/CNI+ | CE/CNI- | CE/CNI+ |
| Total number of samples | N = 14 | N = 13 | N = 14 | N = 13 |
| BGC at D0 (%) - Median | 6.0 | 12.1 | 16.5 | 22.7 |
| BGC at D0 (%) - Range | 3.9-23.3 | 2.5-36.3 | 4.0-63.6 | 8.3-79.5 |
| Number of paired samples | N = 12 | N = 12 | N = 11 | N = 11 |
| BGC at D0 (%) - Median | 6.0 | 12.7 | 16.5 | 22.7 |
| BGC at D0 (%) - Range | 3.9-23.3 | 2.5-36.3 | 4.0-63.6 | 8.3-79.5 |
| *p* value | *p = 0.0121* | | *p = 0.1095* | |

**Supplemental Table 1. Analysis of Biopsy GIC content (BGC) at day 0 (D0) across the different biopsy subgroups.** Statistical analyses were conducted on paired samples.

| | Biopsy localization | | | |
|---|---|---|---|---|
| | FLAIR/CNI- | FLAIR/CNI+ | CE/CNI- | CE/CNI+ |
| Total number of samples | N = 14 | N = 13 | N = 14 | N = 13 |
| Primary NS generation (Y/N) | | | | |
| No | 4 (30.8%) | 2 (15.4%) | 2 (14.3%) | 2 (15.4%) |
| Yes | 9 (69.2%) | 11 (84.6%) | 12 (85.7%) | 11 (84.6%) |
| Missing | 1 | 0 | 0 | 0 |
| If Yes, Time to NS generation (days) | | | | |
| TTN - Median | 21.0 | 13.0 | 9.5 | 8.0 |
| TTN - Range | 8.0-60.0 | 4.0-21.0 | 2.0-50.0 | 1.0-53.0 |
| Secondary NS generation (Y/N) | | | | |
| No | 8 (57.1%) | 4 (30.8%) | 4 (28.6%) | 4 (30.8%) |
| Yes | 6 (42.9%) | 9 (69.2%) | 10 (71.4%) | 9 (69.2%) |
| Number of paired samples | N = 12 | N = 12 | N = 11 | N = 11 |
| Primary NS generation (Y/N) | | | | |
| No | 4 (36.4%) | 2 (16.7%) | 2 (18.2%) | 2 (18.2%) |
| Yes | 7 (63.6%) | 10 (82.2%) | 9 (81.8%) | 9 (81.8%) |
| Missing | 1 | 0 | 0 | 0 |
| *p* value | *p = 0.500* | | *p = 1.000* | |
| If Yes, Time to NS generation (days) | | | | |
| TTN - Median | 18.0 | 13.0 | 11.0 | 11.0 |
| TTN - Range | 8.0-60.0 | 4.0-21.0 | 2.0-50.0 | 2.0-53.0 |
| *p* value | *p = 0.1020* | | *p = 0.6634* | |
| If Yes, Primary NS generation at 14 days (Y/N) | | | | |
| No | 8 (72.7%) | 6 (50.0%) | 5 (45.5%) | 6 (54.5%) |
| Yes | 3 (27.3%) | 6 (50.0%) | 6 (54.5%) | 5 (45.5%) |
| Missing | 1 | 0 | 0 | 0 |
| *p* value | *p = 0.2500* | | *p = 1.000* | |
| If Yes, Primary NS generation at 21 days (Y/N) | | | | |
| No | 6 (54.5%) | 2 (16.7%) | 3 (27.3%) | 4 (36.4%) |
| Yes | 5 (45.5%) | 10 (83.3%) | 8 (72.7%) | 7 (63.6%) |
| Missing | 1 | 0 | 0 | 0 |
| *p* value | *p = 0.1250* | | *p = 1.000* | |
| If Yes, Secondary NS generation (Y/N) | | | | |
| No | 7 (58.3%) | 4 (33.3%) | 3 (27.3%) | 4 (36.4%) |
| Yes | 5 (41.7%) | 8 (66.7%) | 8 (72.7%) | 7 (63.6%) |
| *p* value | *p = 0.2500* | | *p = 1.000* | |

**Supplemental Table 2. Analysis of both ability and delay to generate neurosphere (NS) from biopsies across the different subgroups.** Were evaluated the ability to generate primary or secondary NS (in %), the ability to generate primary NS at day 14 or 21 (in %) and the time to primary NS (TTN) generation from biopsies. Statistical analyses were conducted on paired samples.

| | Univariable analysis of mice OS (Logrank test) | | | |
|---|---|---|---|---|
| | Evt / N | survival at t=250 days | [95%CI] | p-value |
| Type of sample | | | | |
| FLAIR/CNI- | 28/28 | 42.9% | [24.6;60.0] | |
| FLAIR/CNI+ | 40/40 | 17.5% | [7.7;30.6] | *p = 0.0448* |
| Type of sample | | | | |
| CE/CNI- | 40/40 | 20.0% | [9.4;33.5] | |
| CE/CNI+ | 39/39 | 35.9% | [21.4;50.6] | *p = 0.2125* |
| Type of sample | | | | |
| CNI- | 68/68 | 29.4% | [19.1;40.4] | |
| CNI+ | 79/79 | 26.6% | [17.4;36.6] | *p = 0.6621* |
| Type of sample | | | | |
| FLAIR | 68/68 | 27.9% | [17.9;38.9] | |
| CE | 79/79 | 27.8% | [18.5;38.0] | *p = 0.7025* |

**Supplemental Table 3. Analysis of the survival rate in xenografted mice across the different subgroups.** For each primary GIC cell lines able to generate stable secondary neurospheres, NS-dissociated cells were subjected to orthotopic xenografts in nude mice. Through Logrank analysis, the survival rates at t= 250 days were evaluated across the different subgroups.

| | Biopsy localization | | | |
|---|---|---|---|---|
| | FLAIR/CNI- | FLAIR/CNI+ | CE/CNI- | CE/CNI+ |
| Number of paired samples | N = 12 | N = 12 | N = 11 | N = 11 |
| Primary NS generation (Y/N) | | | | |
| No | 4 (36.4%) | 2 (16.7%) | 2 (18.2%) | 2 (18.2%) |
| Yes | 7 (63.6%) | 10 (82.2%) | 9 (81.8%) | 9 (81.8%) |
| Missing | 1 | 0 | 0 | 0 |
| If Yes, MID for radioresistance estimation (Gy) | | | | |
| MID - Median | 4.2 | 3.3 | 4.3 | 4.5 |
| MID - Range | 2.9-7.0 | 2.1-5.1 | 2.8-8.7 | 3.1-6.3 |
| *p* value | *p = 0.4652* | | *p = 1.000* | |

**Supplemental Table 4. Evaluation of the radioresistance capacity for all stable NS cell lines generated from biopsies across the different subgroups.** The in vitro radiosensitivity of each stable NS cell line was studied by realizing clonogenic assays at increasing irradiation doses. The MID (mean inactivation dose) was then established for all cell lines in the four sample subgroups (FLAIR/CNI-; FLAIR/CNI+; CE/CNI-; CE/CNI+). Statistical analyses were conducted on paired samples.

**A**

| | Univariable analysis of OS - BGC% at D0 Continuous variables (Cox model) | | | |
|---|---|---|---|---|
| | **N** | **HR** | **[95%CI]** | **p-value** |
| Flair and CE (CNI-/+) | | | | |
| BGC% maximum | 16 | 1.00 | [0.98-1.03] | *p = 0.848* |
| Flair (CNI-/+) | | | | |
| BGC% maximum in FLAIR | 15 | 1.16 | [1.04-1.29] | ***p = 0.008*** |
| Flair CNI+ | | | | |
| BGC% | 13 | 1.15 | [1.03-1.28] | ***p = 0.010*** |
| CE (CNI-/+) | | | | |
| BGC% maximum in CE | 16 | 1.00 | [0.97-1.02] | *p = 0.960* |

| | Univariable analysis of OS - BGC% at D0 Qualitative variables (Logrank test) | | | | | |
|---|---|---|---|---|---|---|
| | **Evt / N** | **Median** | **[95%CI]** | **HR** | **[95%CI]** | **p-value** |
| Flair and CE (CNI-/+) | | | | | | |
| BGC% maximum | | | | | | |
| ⩽ median | 8 / 8 | 14.3 | [2.8-29.3] | 1.00 | | *p = 0.9486* |
| > median | 8 / 8 | 20.4 | [6.9-27.3] | 1.03 | [0.37-2.89] | |
| Flair (CNI-/+) | | | | | | |
| BGC% maximum in FLAIR | | | | | | |
| ⩽ median | 8 / 8 | 21.1 | [8.0-27.3] | 1.00 | | *p = 0.1484* |
| > median | 7 / 7 | 14.3 | [2.8-20.1] | 2.18 | [0.74-6.45] | |
| Flair CNI+ | | | | | | |
| BGC% | | | | | | |
| ⩽ median | 7 / 7 | 25.7 | [8.0-30.7] | 1.00 | | ***p = 0.0112*** |
| > median | 6 / 6 | 12.7 | [2.8- . ] | 6.47 | [1.26-33.11] | |
| CE (CNI-/+) | | | | | | |
| BGC% maximum in CE | | | | | | |
| ⩽ median | 8 / 8 | 14.3 | [2.8-29.3] | 1.00 | | *p = 0.9486* |
| > median | 8 / 8 | 20.4 | [6.9-27.3] | 1.03 | [0.37-2.89] | |

**B**

| | Univariable analysis of PFS - BGC% at D0 | | | |
|---|---|---|---|---|
| | **N** | **HR** | **[95%CI]** | **p-value** |
| Flair and CE (CNI-/+) | | | | |
| BGC% maximum | 16 | 1.00 | [0.98-1.03] | *p = 0.724* |
| Flair (CNI-/+) | | | | |
| BGC% maximum in FLAIR | 15 | 1.14 | [1.04-1.24] | ***p = 0.004*** |
| Flair CNI+ | | | | |
| BGC% | 13 | 1.16 | [1.04-1.30] | ***p = 0.006*** |
| CE (CNI-/+) | | | | |
| BGC% maximum in CE | 16 | 1.00 | [0.98-1.03] | *p = 0.908* |

| | Univariable analysis of PFS - BGC% at D0 Qualitative variables (Logrank test) | | | | | |
|---|---|---|---|---|---|---|
| | **Evt / N** | **Median** | **[95%CI]** | **HR** | **[95%CI]** | **p-value** |
| Flair and CE (CNI-/+) | | | | | | |
| BGC% maximum | | | | | | |
| ⩽ median | 8 / 8 | 6.5 | [2.8-12.5] | 1.00 | | *p = 0.9157* |
| > median | 8 / 8 | 6.1 | [3.7-15.1] | 1.06 | [0.38-2.97] | |
| Flair (CNI-/+) | | | | | | |
| BGC% maximum in FLAIR | | | | | | |
| ⩽ median | 8 / 8 | 9.1 | [3.7-21.1] | 1.00 | | *p = 0.0657* |
| > median | 7 / 7 | 5.6 | [2.8-8.7] | 2.89 | [0.89-9.37] | |
| Flair CNI+ | | | | | | |
| BGC% | | | | | | |
| ⩽ median | 7 / 7 | 10.7 | [6.1-15.1] | 1.00 | | ***p = 0.0057*** |
| > median | 6 / 6 | 5.5 | [2.8- .] | 7.36 | [1.44-37.57] | |
| CE (CNI-/+) | | | | | | |
| BGC% maximum in CE | | | | | | |
| ⩽ median | 8 / 8 | 6.5 | [2.8-12.5] | 1.00 | | *p = 0.9157* |
| > median | 8 / 8 | 6.1 | [3.7-15.1] | 1.06 | [0.38-2.97] | |

**Supplemental Table 5. Relationship between Biopsy GIC content (BGC) at day 0 (D0) in specified biopsy subgroups and patient outcome.** The relationship between BGC% maximal value in the indicated sample groups (all FLAIR+CE samples, all FLAIR samples, all CE samples or only FLAIR/CNI+ samples) and either patient Overall Survival (OS) **(A)** or Progression Free Survival (PFS) **(B)** was studied by univariable analyses using either continuous variables (Cox model, upper panels) or qualitative variable (Logrank test, lower panels). HR: Hazard Ratio

**A**

| | Univariable analysis of OS - TTN Continuous variables (Cox model) | | | |
|---|---|---|---|---|
| | **N** | **HR** | **[95%CI]** | **p-value** |
| Flair and CE (CNI-/+) | | | | |
| TTN (days) minimum | 16 | 1.01 | [0.95-1.07] | *p = 0.779* |
| Flair (CNI-/+) | | | | |
| TTN (days) minimum in FLAIR | 13 | 0.97 | [0.91-1.04] | *p = 0.415* |
| Flair CNI+ | | | | |
| TTN (days) | 11 | 0.95 | [0.84-1.07] | *p = 0.385* |
| CE (CNI-/+) | | | | |
| TTN (days) minimum in CE | 15 | 1.01 | [0.95-1.08] | *p = 0.707* |

**B**

| | Univariable analysis of PFS - TTN Continuous variables (Cox model) | | | |
|---|---|---|---|---|
| | **N** | **HR** | **[95%CI]** | **p-value** |
| Flair and CE (CNI-/+) | | | | |
| TTN (days) minimum | 16 | 0.98 | [0.93-1.04] | *p = 0.542* |
| Flair (CNI-/+) | | | | |
| TTN (days) minimum in FLAIR | 13 | 0.97 | [0.90-1.05] | *p = 0.518* |
| Flair CNI+ | | | | |
| TTN (days) | 11 | 0.94 | [0.81-1.08] | *p = 0.386* |
| CE (CNI-/+) | | | | |
| TTN (days) minimum in CE | 15 | 0.99 | [0.93-1.04] | *p = 0.605* |

**Supplemental Table 6. Relationship between Time to NS generation (TTN) in specified biopsy subgroups and patient outcome.** The relationship between TTN minimal value in the indicated sample groups (all FLAIR+CE samples, all FLAIR samples, all CE samples or only FLAIR/CNI+ samples) and either patient Overall Survival (OS) **(A)** or Progression Free Survival (PFS) **(B)** was studied by univariable analysis using continuous variables (Cox model). HR: Hazard Ratio

**A**

| | Univariable analysis of OS - TTN Qualitative variables (Logrank test) | | | | | |
|---|---|---|---|---|---|---|
| | **Evt / N** | **Median** | **[95%CI]** | **HR** | **[95%CI]** | **p-value** |
| Flair (CNI-/+) | | | | | | |
| Primary NS generation at 14 days in FLAIR (Y/N) | | | | | | |
| No | 8 / 8 | 21.1 | [17.0-30.7] | 1.00 | | ***p = 0.0147*** |
| Yes | 7 / 7 | 12.7 | [2.8-20.1] | 3.96 | [1.22-12.89] | |
| Primary NS generation at 21 days in FLAIR (Y/N) | | | | | | |
| No | 4 / 4 | 21.1 | [20.4- .] | 1.00 | | *p = 0.5776* |
| Yes | 11 / 11 | 17.0 | [6.9-25.7] | 1.40 | [0.42-4.66] | |
| Flair CNI+ | | | | | | |
| Primary NS generation at 14 days (Y/N) | | | | | | |
| No | 6 / 6 | 20.4 | [17.0- .] | 1.00 | | ***p = 0.0190*** |
| Yes | 7 / 7 | 12.7 | [2.8-20.1] | 4.60 | [1.15-18.37] | |
| CE (CNI-/+) | | | | | | |
| Primary NS generation at 14 days in CE (Y/N) | | | | | | |
| No | 4 / 4 | 14.3 | [2.8- .] | 1.00 | | *p = 0.8721* |
| Yes | 12 / 12 | 20.1 | [8.0-27.3] | 0.91 | [0.28-2.91] | |

**B**

| | Univariable analysis of PFS - TTN Qualitative variables (Logrank test) | | | | | |
|---|---|---|---|---|---|---|
| | **Evt / N** | **Median** | **[95%CI]** | **HR** | **[95%CI]** | **p-value** |
| Flair (CNI-/+) | | | | | | |
| Primary NS generation at 14 days in FLAIR (Y/N) | | | | | | |
| No | 8 / 8 | 9.1 | [3.7-21.1] | 1.00 | | *p = 0.0643* |
| Yes | 7 / 7 | 6.2 | [2.8-8.7] | 3.13 | [0.88-11.11] | |
| Primary NS generation at 21 days in FLAIR (Y/N) | | | | | | |
| No | 4 / 4 | 9.1 | [3.7- .] | 1.00 | | *p = 0.4561* |
| Yes | 11 / 11 | 6.2 | [5.2-10.7] | 1.57 | [0.48-5.19] | |
| Flair CNI+ | | | | | | |
| Primary NS generation at 14 days (Y/N) | | | | | | |
| No | 6 / 6 | 9.1 | [5.5- .] | 1.00 | | *p = 0.0606* |
| Yes | 7 / 7 | 6.2 | [2.8-8.7] | 3.53 | [0.88-14.19] | |
| CE (CNI-/+) | | | | | | |
| Primary NS generation at 14 days in CE (Y/N) | | | | | | |
| No | 4 / 4 | 6.5 | [2.8- .] | 1.00 | | *p = 0.7100* |
| Yes | 12 / 12 | 6.2 | [5.2-12.5] | 1.25 | [0.39-4.00] | |

**Supplemental Table 7. Relationship between the ability to generate primary NS at day 14 or day 21 in specified biopsy subgroups and patient outcome.** The relationship between the capacity to generate primary NS at day 14 (or day 21) in the indicated sample groups (all FLAIR samples, all CE samples or only FLAIR/CNI+ samples) and either patient Overall Survival (OS) **(A)** or Progression Free Survival (PFS) **(B)** was studied by univariable analysis using qualitative variable (Logrank test). HR: Hazard Ratio

| | Gene Signatures | | | |
|---|---|---|---|---|
| | Astrocytic | Neuronal | Oligodendrocytic | Stemness |
| Gene names | GFAP | NCAM1 | CLDN11 | ITGB8 |
| | GLUL | MAP2 | MYRF | NOTCH2 |
| | SLC1A2 | STMN1 | MOG | NES |
| | AQP4 | SNCA | SOX10 | STAT3 |
| | SLC1A3 | SYP | OMG | NOTCH1 |
| | NDRG2 | NEFL | MAL | ITGA6 |
| | S100B | SLC17A7 | OPALIN | SOX2 |
| | GJA1 | SYN1 | | CD44 |
| | | NEFH | | NOTCH3 |
| | | NEFM | | CXCR4 |
| | | RBFOX3 | | CSPG4 |
| | | | | BMI1 |
| | | | | NOTCH4 |
| | | | | EPHB2 |
| | | | | MYC |
| | | | | PROM1 |
| | | | | MET |
| | | | | EPHA2 |
| | | | | BIRC5 |
| | | | | FUT4 |
| | | | | EZH2 |
| | | | | MSI1 |
| | | | | L1CAM |
| | | | | SOX1 |
| | | | | SALL4 |
| **Number of genes** | 8 | 11 | 7 | 25 |

**Supplemental Table 8. Custom gene signatures used for enrichment analyses performed on RNAseq data obtained from patient tumor biopsies.**
